# Mapping the genetic landscape of disorders on the autoimmune-autoinflammatory continuum: potential implications for classification and treatment

**DOI:** 10.1101/2024.08.08.24311518

**Authors:** Vera Fominykh, Alexey A. Shadrin, Piotr Jaholkowski, Julian Fuhrer, Nadine Parker, Erik D. Wiström, Oleksandr Frei, Olav B. Smeland, Helga Sanner, Srdjan Djurovic, Ole A. Andreassen

## Abstract

**Objectives:** Based on clinical, biomarker and genetic data, autoimmune and autoinflammatory disorders (AIDs) can be classified as a disease continuum from pure autoinflammatory to pure autoimmune with mixed diseases in between. However, the genetic architecture of AIDs has not been systematically described. Here we investigate the polygenic continuum of AIDs using genome-wide association studies (GWAS) and statistical genetics methods.

**Methods:** We mapped the genetic landscape of 15 AIDs using GWAS summary statistics and methods including genomic structural equation modelling (genomic SEM), linkage disequilibrium score regression, Local Analysis of [co]Variant Association, and Gaussian causal mixture modelling (MiXeR). We performed enrichment analyses of tissues and biological gene-sets using MAGMA.

**Results:** Genomic SEM suggested a continuum structure with four underlying latent factors from autoimmune diseases at one end to autoinflammatory on the opposite end. Across AIDs, we observed a balanced mixture of negative and positive local correlations within the major histocompatibility complex, while outside this region they were predominantly positive. MiXeR analysis showed large genetic overlap in accordance with the continuum landscape. MAGMA analysis implicated genes associated with monogenic immune diseases in autoimmune (factor 1) and autoinflammatory diseases (factor 4).

**Conclusions:** Our findings support a polygenic continuum across AIDs, with four genetic clusters. The “polygenic autoimmune” and “polygenic autoinflammatory” clusters reside on margins of this continuum. The identified genetic patterns across different AIDs can potentially guide drug selection, as patients within the same clusters may benefit from the same therapies.

## Introduction

During the last twenty years, autoimmune and autoinflammatory disorders (AIDs) have entered the frontline of the clinical field and pharmaceutical development in neurology and rheumatology [Mukai et al., 2023, Mueller et al., 2023]. Various new diseases have been described, including autoimmune encephalitis [Dalmau et al., 2018], myelin oligodendrocyte glycoprotein-associated diseases, monogenic autoinflammatory diseases (interferonopathies, adenosine deaminase 2 deficiency) and others [Canna et al., 2015, Di Donato et al., 2021, de Jesus et al., 2023]. The global rise in AIDs, as highlighted by epidemiological studies [Conrad et al., 2023, Miller et al, 2023], has sparked a need for urgent innovation in the development of curation approaches and treatment strategies.

In addition, the release of new therapeutic options for neuromyelitis optica [Held et al., 2021], myasthenia gravis [DeHart-McCoyle et al., 2022] and monogenic immune diseases [Perez et al., 2022] bring new attention of clinicians to immune disease conception, classification and genetics. The current understanding of AIDs relies on previous immunological concepts of autoinflammation and autoimmunity, which define autoinflammation as a dysregulated activation of innate immune cells, driven by an imbalance in the axis of pro-inflammatory cytokines, which leads to damage of host tissues without a break in immune tolerance [Savic et al., 2020, Szekanecz et al., 2021]. Conversely, autoimmunity is characterised by the loss of immune tolerance, the recognition of self-antigens and the activation of T cells and B cells, followed by the production of specific autoantibodies and the damage of multiple organs owing to a dysregulated adaptive immune response [McGonagle et al., 2006, Matzinger et al., 2022, Shirafkan et al, 2024]. Regarding the fact that AIDs is spread between different medical specialities (neurology, rheumatology, endocrinology, gastroenterology etc) the new methods and data driven approach can help with improving classification which is needed.

The conceptualization of AIDs as a continuum began with proof of concept publications which suggested the classification of immunological diseases based on clinical and laboratory data [McGonagle et al. 2006 updated by Ben-Chetrit et al, 2018 and Savic et al, 2020], and described AIDs as a continuum with 5 main classes (monogenic autoinflammatory, polygenic autoinflammatory, mixed pattern, polygenic autoimmune, monogenic autoimmune diseases) highlighting the importance of genetic factors for the classification [See Fig 1.]. This classification has become commonplace in rheumatology, where SLE represents the prototype of systemic autoimmunity with production of multiple autoantibodies. There is still no consensus on the precise classification of some diseases such as rheumatoid arthritis (RA), juvenile idiopathic arthritis (JIA) and ankylosing spondylitis (AS) [Mauro et al., 2021], since they exhibit overlapping features of both autoimmunity and autoinflammation [Szekanecz et al., 2021]. In neurology some diseases are easier to place on the AIDs continuum (for example, myasthenia or anti-LGi1-positive autoimmune encephalitis are considered to be classical autoimmune diseases [Fichtner et al., 2020, Bink et al., 2017] but for the majority it remains unclear (for example, central nervous system vasculitis, acute encephalomyelitis, Höftberger et al., 2017, de Moraes et al., 2023]. Moreover, current efforts are based on clinical features and qualitative markers, leading to competing classifications due to different hierarchical structures of clinical terms and biomarkers [Grateu et al., 2013, Pathak et al., 2017].

**Fig. 1.**
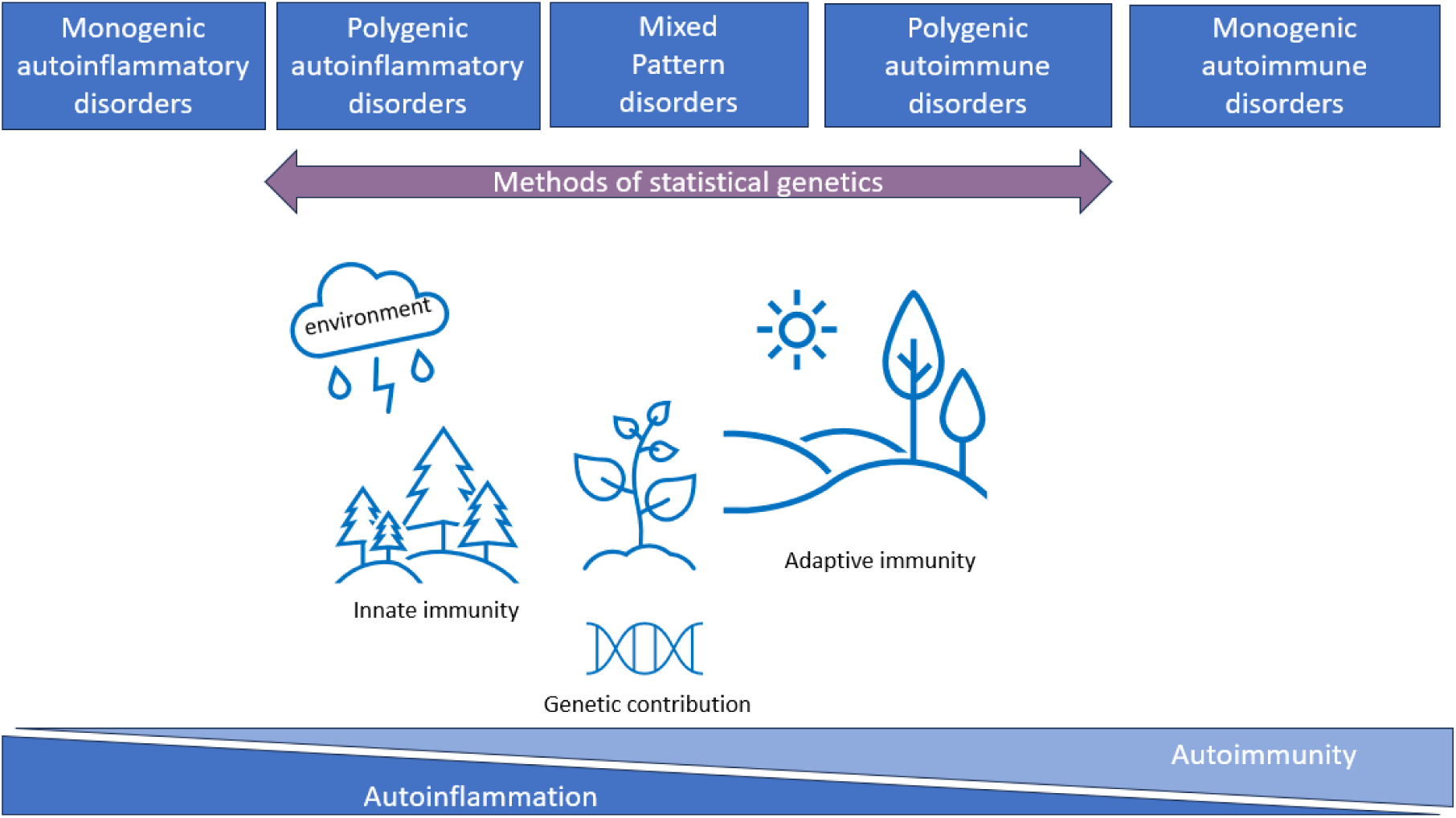
The immunological disease landscape. AIDs described as a continuum with five main classes (monogenic autoinflammatory, polygenic autoinflammatory, mixed pattern, polygenic autoimmune, monogenic autoimmune disorders) highlighting the importance of genetic factors for the classification. Methods of statistical genetics can work with polygenic disorders and can help to characterise this continuum from genetics perspectives using available GWAS data.

AIDs have a high degree of comorbidity within families [Harroud et al., 2023, Barkhane et al., 2022] and twins [Gerussi et al., 2022] suggesting shared genetic risk factors. Additionally, the co-existence of them in one individual suggests shared genetic underpinnings across different immune pathologies [Remalante-Rayco et al., 2023, Fominykh et al., 2017]. Genetic-based disease classification has been used in a study from Japan Biobank [Sakaue et al., 2021] and a cross-disorder genetic analysis of immune disease [Demela et al., 2023, Lincoln et al., 2024, Topaloudi et al., 2023]. Nevertheless, most of the available cross-disorder genetic studies have used only a few disorders, without clearly defined selection, and focusing on a shared component but not on the characterising of separate clusters. Only a limited number of studies have systematically evaluated the AIDs continuum. A great effort was made to use the AutoCore network for the integration of a set of 186 inborn errors of immunity [Bousfiha et al., 2020] with predominant autoimmunity or autoinflammation into a comprehensive map of human immune dysregulation [Guthrie et al., 2023]. In France, the TransImmunome project was started in 2015 to revisit the nosology of AIDs by combining clinical and biomarkers information [Lorenzon et al., 2018, Tchitchek et al., 2024]. State-of-art methods of statistical genetics provide an additional opportunity to address this heterogeneity between AIDs through classifying diseases into more homogeneous subgroups based on the underlying genes and pathways that drive disease.

The main aims of our study are a) to evaluate the hypothesis of AIDs continuum based on GWAS data and b) to identify genetically driven clusters of closely related disorders and their shared genetic factors using publicly available large datasets and state-of-the art methodology. Bridging genetically driven classification with existing clinical knowledge may provide a better understanding of disease-specific molecular mechanisms, guiding further investigations of AIDs and facilitating identification of potential targets for drug repurposing. The results can help to raise awareness of the distinct pathophysiology of AIDs, and to identify potential research directions in the field of immune-mediated disorders.

## 2 Materials and methods

### 2.1. Samples

We curated a collection of well-powered publicly available GWAS summary statistics which belong to neurological, rheumatological, gastroenterological, and endocrine system AIDs resulting in data on 15 immune-linked diseases (Table 1). The cut-off for GWAS inclusion was August 2023. First, we have 26 immune-linked disorders in the list (Suppl.1, Table 1) with existing GWASs and applied criteria with N effective sample size greater than 5000 [Bulik-Sullivan et al., 2015, Tylee et al., 2018] and excluded datasets with less than 200,000 SNPs overlap with the linkage disequilibrium (LD) score regression reference panel [Zheng et al., 2017]. N effective was computed with the formula 4/(1/n cases + 1/n controls) or for internal meta-analysis (described below) as the sum of N effective for contributing cohorts. This resulted in seventeen diseases. However, we had to exclude ankylosing spondylitis and narcolepsy because only ImmunoChip data was available which didn’t provide genome-wide coverage. For SLE, JIA, PS and CeD we performed in-house meta-analyses using METAL from different publicly available datasets (FinnGen R9 release and available published GWAS, the source of data mentioned in Table 1 and Suppl. 1, Table 2) in order to increase power of current available GWASes. As well we can highlight the need for more powerful GWASes in immune-mediated neurological diseases because we can keep only two among neurological diseases. All GWAS data were limited to participants of European ancestry. After data harmonisation and pre-processing of the GWAS summary statistics with PythonConvert (https://github.com/precimed/python_convert/), we conducted cross-trait analyses using a variety of analytical tools as described below.

**Table 1.** Overview of the GWAS used in the study.

|  | Diseases | Abbreviation | N cases | N controls | N effective | Source of GWAS (PMID/ GWAS catalog) |
| --- | --- | --- | --- | --- | --- | --- |
| 1 | Autoimmune thyroiditis | AITD | 30234 | 725172 | 114296 | 32581359 |
| 2 | Celiac disease | CeD | 6897 | 334824 | 22142 | 20190752, 34278373 |
| 3 | Crohn's disease | CD | 12194 | 34915 | 36151 | 28067908 |
| 4 | Juvenile idiopathic arthritis | JIA | 4799 | 294231 | 15670 | FinnGen, 33106285 |
| 5 | Multiple sclerosis | MS | 14802 | 26703 | 38093 | 31604244 |
| 6 | Myasthenia gravis | MG | 1873 | 36370 | 7125 | 35074870 |
| 7 | Primary biliary cholangitis | PBC | 8021 | 16489 | 21584 | 34033851 |
| 8 | Primary sclerosing cholangitis | PSC | 2 871 | 12019 | 9270 | 27992413 |
| 9 | Primary Sjogren's syndrome | SjS | 3232 | 17481 | 10911 | 35896530 |
| 10 | Psoriasis (including PsA) | PS | 17255 | 693100 | 64320 | FinnGen, 34278373, 24482804 |
| 11 | Rheumatoid arthritis | RA | 22350 | 74823 | 68838 | GCST 90132223 |
| 12 | Systemic lupus erythematosus | SLE | 5595 | 361571 | 15096 | FinnGen, 26502338, 29848360 |
| 13 | Systemic sclerosis | SS | 9095 | 17584 | 23978 | GCST 31672989 |
| 14 | Type 1 diabetes | T1D | 18 942 | 501 638 | 73011 | 34012112 |
| 15 | Ulcerative colitis | UC | 12366 | 34915 | 36527 | 28067908 |

**Table 2.** genomic SEM model performance criteria: AIC — Akaike Information Criterion, CFI — Comparative Fit Index, SRMR — standardised root mean square residual, χ^2^ — model chi-square, reflecting index of exact fit to observed data df and p_χ2_ — degrees of freedom and p-value for the model χ^2^.

| **N factors** | **$\chi^2$** | **df** | **$p\chi^2$** | **AIC** | **CFI** | **SRMR** |
| --- | --- | --- | --- | --- | --- | --- |
| 1 | 1321.83 | 90 | 4.56E-218 | 1381.83 | 0.658 | 0.124 |
| 2 | 1073.7 | 88 | 3.08E-169 | 1137.69 | 0.727 | 0.111 |
| 3 | 779.35 | 87 | 9.09E-112 | 845.35 | 0.808 | 0.094 |
| **4** | **215.59** | **56** | **1.42E-20** | **285.59** | **0.919** | **0.077** |
| 5 | 238.37 | 54 | 5.25E-25 | 312.37 | 0.907 | 0.083 |

### 2.2 Analytical tools

#### 2.2.1 Genomic structural equation modelling (SEM)

Genomic SEM [Grotzinger et al., 2019] models the multivariate genetic architecture across traits and may reveal latent factors underlying genetic correlations and clusters of correlated traits and determine how latent factors correlate with each other. We conducted exploratory and confirmatory factor analyses (EFA and CFA, respectively) of the 15 immune phenotypes. First, the multivariable extension of LD score regression (LDSC) employed in genomic SEM was used to derive a genetic covariance matrix (S) and sampling covariance matrix (V). Population prevalence was taken from the literature (Suppl.1, Table 3). Next, EFA with promax rotation was conducted on the standardised S matrix using the R genomic SEM package (R version: R 4.3.2, https://github.com/GenomicSEM/, Suppl 1., Table 4). Results from the EFA were used to guide CFA for a one-, two-, three-, four- and five-factor model. CFA was performed using Genomic SEM, and correlated factors with standardised loadings > 0.25 were retained for CFA. Model fit (Table 2 and Suppl. 1, Table 5) for each factor model was assessed using recommended fit indices: standardised root mean square residual (SRMR), model χ^2^ statistic, Akaike Information Criterion (AIC), and Comparative Fit Index (CFI). Model fit was considered acceptable for CFI values ≥ 0.90 and SRMR < 0.1 [for interpretation of model fit Grotzinger et al., 2019].

#### 2.2.2 Linkage disequilibrium score regression (LDSC) and Gaussian causal mixture modelling (MiXeR)

For each phenotype, we estimated the SNP-heritability using LDSC and used LDSC for establishing the genome-wide genetic correlation (rg) [Bulik-Sullivan et al., 2015]. We used MiXeR (univariate and bivariate, Frei et al., 2019, https://github.com/precimed/mixer) to estimate the number of variants influencing the trait and the number of variants shared between pairs of traits (trait-specific and shared polygenicity) as well as mean effect size across trait influencing variants (discoverability). For MiXeR analyses we excluded the MHC region as described by Frei et al. 2019 (MHC region, 26-34 Mb), due to the intricate LD structure. For assessment of model robustness, delta AIC >0 and visual evaluation of log-likelihood plots were used. Results are presented in heatmap (Fig. 3) showing the proportion of trait-specific and shared trait-influencing SNPs followed by the standard deviation across 20 independent runs, and log-likelihood plots and tables with parameters estimated by the MiXeR model (Suppl.1., Table 7 and 8). For the analysis of disease triplets, we used trivariate MiXeR [Shadrin at al., 2024, https://codeberg.org/intercm/mix3r] constituting a modification of bivariate MiXeR with the possibility to access overlap between 3 phenotypes simultaneously.

#### 2.2.3. Local Analysis of [co]Variant Association (LAVA)

For local r_g_ analyses, we used Local Analysis of [co]Variant Association (LAVA) version 1.3.8, following the protocol with the LD reference panel based on 1000 Genomes phase 3 genotype data for European samples, and the partition of the genome into 2495 regions with an average size of 1 Mb as described elsewhere [Werme J et al, 2022]. We applied Bonferroni correction to account for multiple comparisons within each pairwise analysis. The statistical tests conducted were all two-sided. Then we identified ten regions with the greatest number of significant local genetic correlations across immune traits and created network plots displaying these associations. Only regions revealing significant estimated SNP heritability (p<0.05/2495) in both diseases were used to estimate local genetic correlations between the traits. Also, we described significantly correlated loci in the MHC region (26-34 Mb) across all phenotypes.

#### 2.2.4. Functional Annotation

We followed the FUMA protocol for the identification of genomic loci and genes, as well as performing MAGMA analysis [Watanabe et al., 2017, de Leeuw et al, 2015]. We applied MAGMA to summary statistics to test for enrichment of GWAS signals in 54 tissues. The 54 gene sets were defined by gene-expression levels from 54 GTEx tissues [GTEx Consortium, 2015]. We also analysed genes revealed by MAGMA in each of the 15 diseases and common for all phenotypes, as well as unique genes for diseases assigned to marginal latent factors obtained in genomic SEM analysis. Additionally, unique gene sets were detected for each factor (we applied Bonferroni correction for each trait and then again for 15 comparisons). We used Cytoscape version 3.9.1 [Shannon P et al 2003] for pathway analysis.

## 3. Results

### 3.1. Genomic SEM

To investigate whether there is a common genetic factor underlying the 15 AIDs, we first estimated genetic correlations using the multivariate LDSC implementation in genomic SEM. Factor is a united genetic entity underlying the set of disorders and combines it to the cluster. Next, we uncovered latent factors which represent shared variance components across diseases, we modelled the genetic variance-covariance matrices across traits using genomic SEM (Fig. 2.). The model with four latent factors provides the best fit among all tested models presented in Table 2. Unfortunately, we were unable to include T1D and MS into the four-factor model as the EFA revealed that both traits had multiple, below threshold loadings across the factors (see 4 factor EFA in Suppl. 1., Table 4), leading to a decreased CFA model fit.

**Fig. 2.**
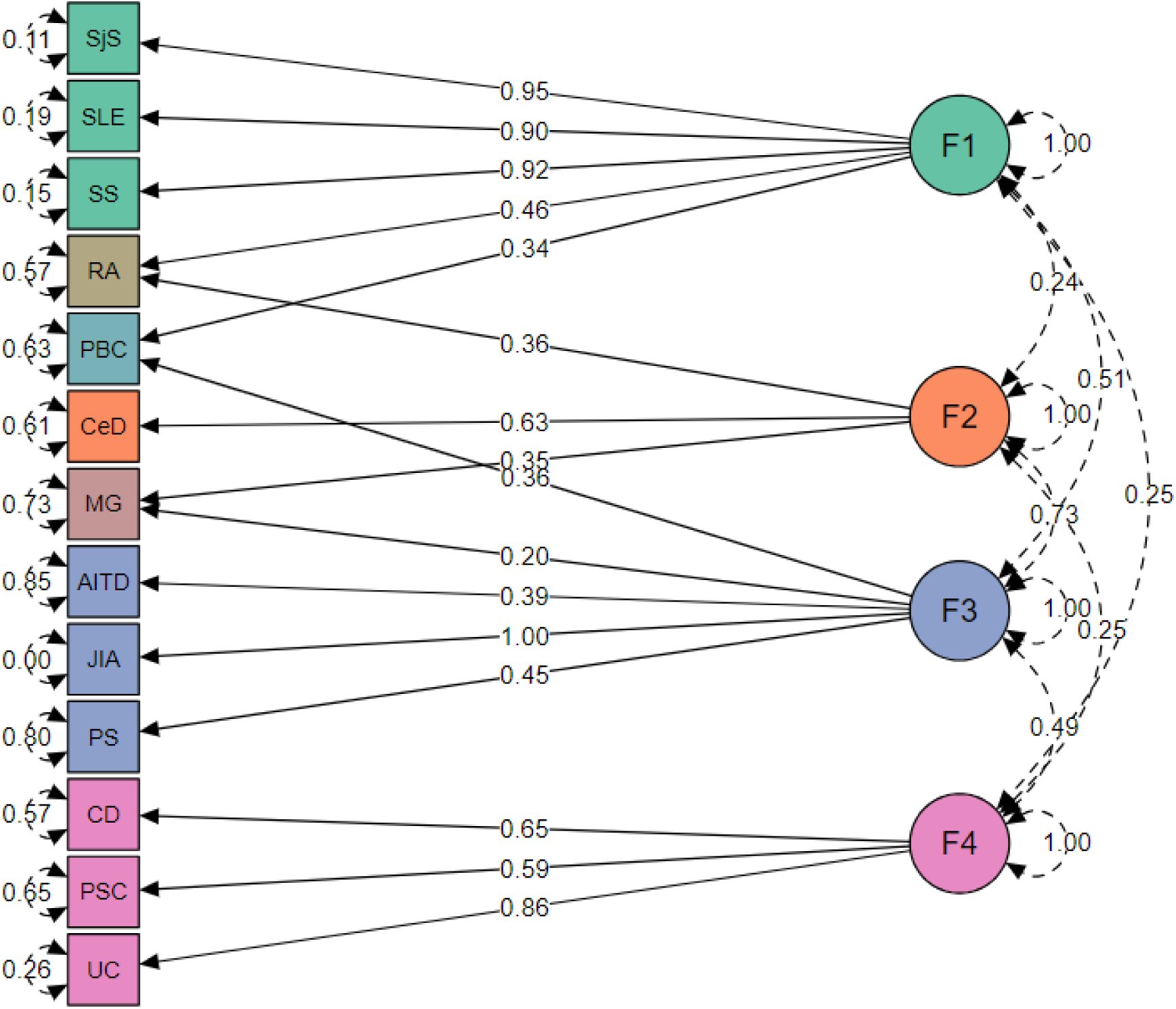
Four groups of immune-mediated diseases identified in genomic SEM analysis. Genomic SEM model with four latent factors representing four diseases clusters. Green represents factor 1 (F1, “autoimmune”), orange factor 2 (F2, “autoimmune-mixed”), blue factor 3 (F3, “mixed pattern”), purple factor 4 (F4, “autoinflammatory”), and diseases cross-load on two factors are shown in its own colour. The arrows connecting the latent variables with diseases are shown “factor loadings” obtained from confirmatory factor analysis. The rounded and dashed arrows on the indicators (traits) are residual variances in the genetic indicators not explained by the common factor. The dashed arrows connecting factors are covariances and give an idea about the factors’ mutual associations.

Factor 1 consists of rheumatic diseases with autoimmune origin (SjS, SLE, SS). Factor 2 consists of MG and CeD which are confirmed to be autoimmune according to the current autoimmune concept, but MG also shared part of genetics with Factor 3, which belongs to mixer pathology. Factor 3 consists of AITD, JIA, PS which are considered as autoimmune, autoinflammatory or mixed according to different classifications. Factor 4 consists of diseases with gastrointestinal tract with autoinflammatory origin (CD, UC and PSC). Several diseases cross-loaded on two factors, such as RA (equally belongs to F1 and F2), PBC (F1 and F3) and MG (F2 and F3). Loadings of T1D and MS were below the selected cutoff for the minimal loading (0.25) and therefore don’t belong to any of these factors.

#### 3.2. LDSC and MiXeR

We ran LDSC analysis to quantify SNP-heritability, observed scale (Fig. 3, and Suppl. 2, Fig. 1), and univariate and bivariate MiXeR for all phenotypes to characterise polygenicity and overlap between diseases. Univariate MiXeR models exhibited good model fit (Suppl 1., Table 7). As shown below, CD, SLE and PBC have a higher SNP-heritability compared to others (more than 0.45). CeD and SjS have the lowest polygenicity, and RA and AITD have the highest polygenicity (Fig. 3). Bivariate analysis with several diseases, including CeD, SjS and PSC as well as pairs of diseases including SLE-SS and RA-SS didn’t fulfil model robustness criteria and therefore are not presented. Bivariate results, for those diseases with acceptable model fit, are presented in Suppl. 2, Fig. 2, and Suppl. 1, Table 8. We ran trivariate MiXeR for F1 and F4 clusters to show that diseases from the same factor have more overlap compared to another factor, which revealed SLE/SS has less overlap with UC compared with RA from the same factor (Fig. 4).

**Fig.3.**
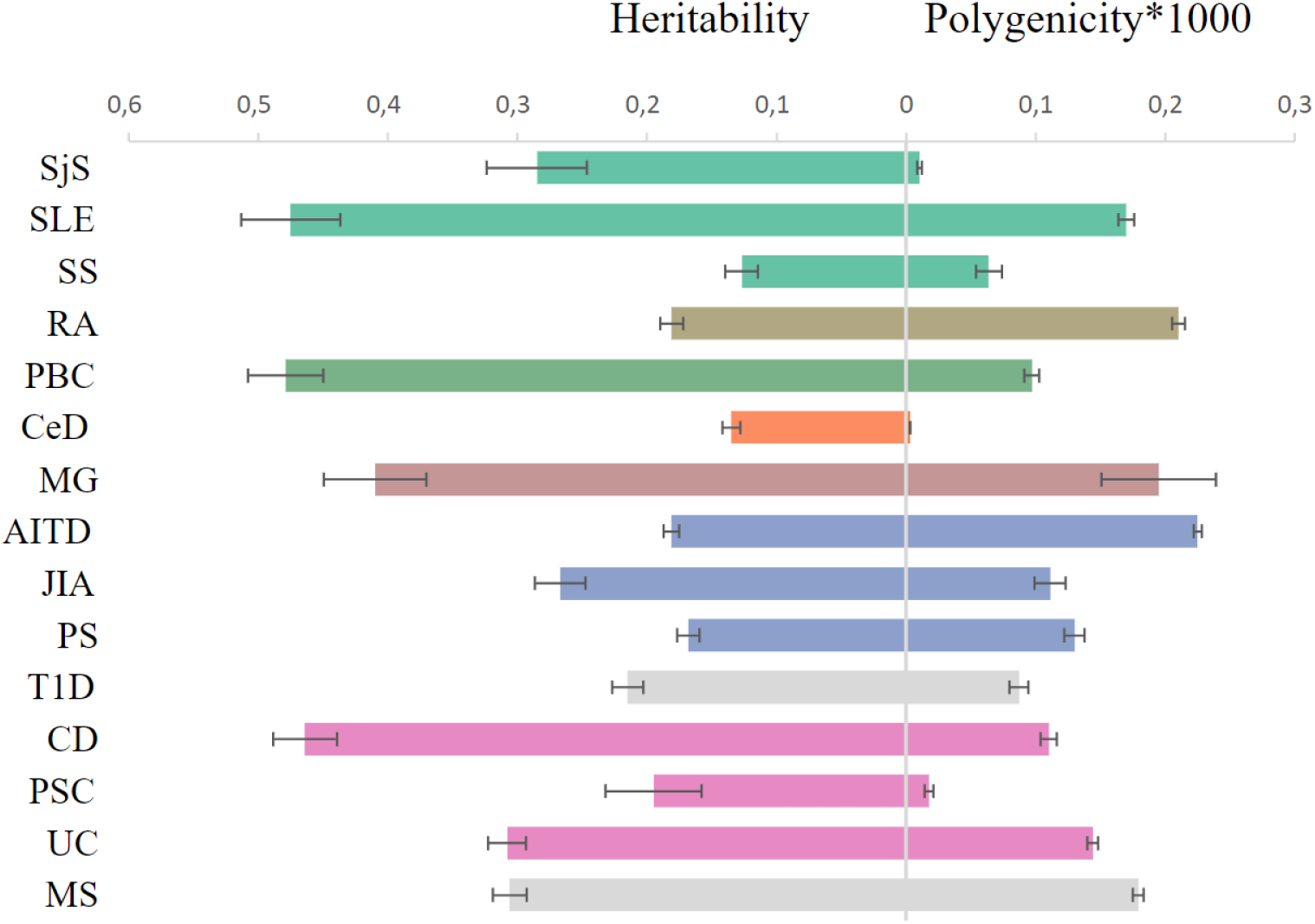
Genetic architecture characteristics. Left part: single-nucleotide polymorphism-based heritability, observed scale, for 15 immune-mediated diseases according to linkage disequilibrium score regression. Right part: the polygenicity value multiplied by 1000 according to Gaussian causal mixture modelling. The colour of dots corresponds to colour in genomic SEM figure (green factor 1, orange factor 2, blue factor 3, purple factor 4 and own colour for mixed diseases).

**Fig.4.**
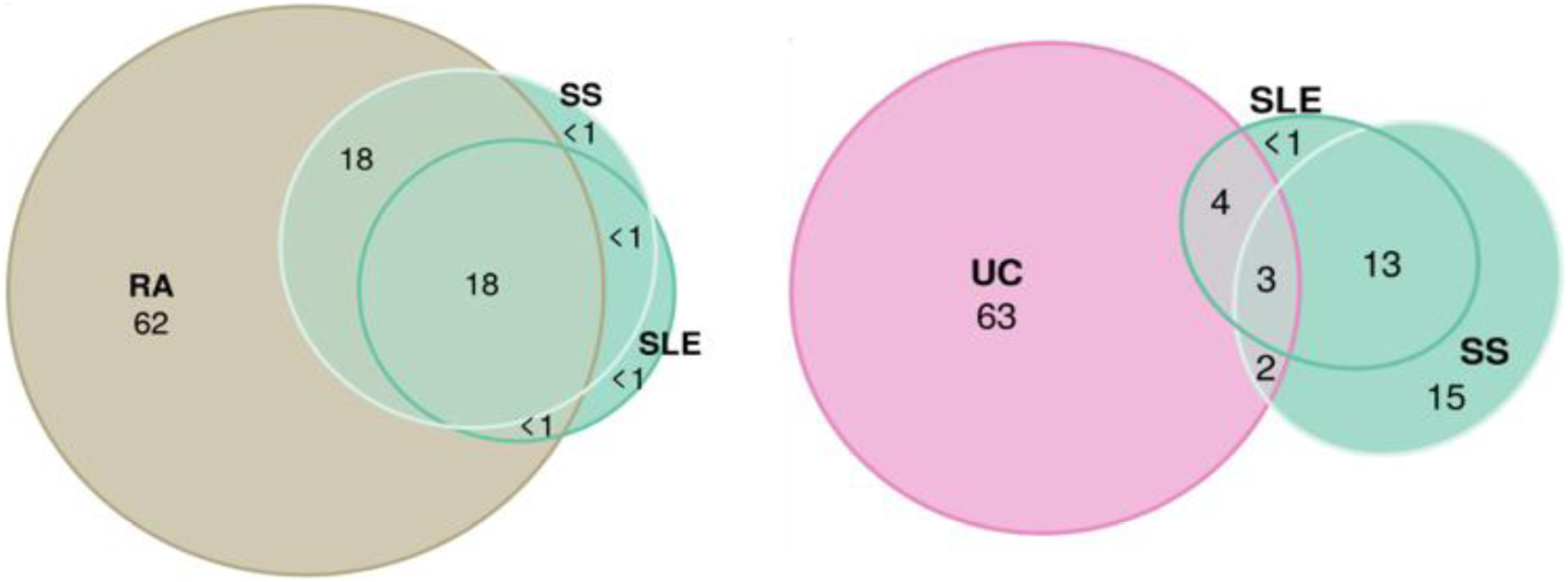
Trivariate Gaussian causal mixture modelling. The illustrations of overlap inside “autoimmune” factor 1 (Systemic Sclerosis, Systemic Lupus Erythematosus and Rheumatoid Arthritis), and between “autoimmune” factor 1 (Systemic Sclerosis, Systemic Lupus Erythematosus) and “autoinflammatory” factor 4 (Ulcerative Colitis). For each triad of phenotypes, for every area of the diagram, its percentage is shown with respect to the combined total area of three phenotypes (rounded to the closest integer). The colour of circles corresponds to colour in genomic SEM figure (green F1, purple F4 and beige for the Rheumatoid Arthritis).

#### 3.3. LAVA across immune-related diseases

Local genetic correlation analyses complemented global genetic correlation between the immune-related phenotypes. This step allowed us to show in which immune disorders genetic correlations are restricted to specific genomic regions, and to identify the shared genetic factors located within these genomic regions.

We identified significant regional correlations between immune diseases and genomic loci, including loci that contain genes known to be implicated in immune processes. SLE, JIA and MG were excluded due to the methods failing to converge. During assessment of the pattern of intercorrelations (Suppl. 2, Fig 3) we revealed that all diseases are highly correlated. We confirmed that the local correlation and heritability were prominent in the MHC region, where strong genetic risk was shown. Among 20 regions with top significant local genetic correlation (see Suppl. 3, Figures and Tables) ten were in the MHC region (26-34 Mb) with the top 1 being in HLA DRB1 gene, top 2 in HLA DQB1 and the top 4-5 in MHC region with different genes. The pattern of local genetic correlation in the MHC region (Fig. 5) involved a mixture of concordant and discordant effects across the diseases. For example, in loci on chr 6 [32539568 - 32586784] UC was negatively correlated with PBC, PSC, PS, AITD and RA but positively correlated with MS. In comparison, fewer diseases displayed significant correlations in loci outside the MHC, but the correlations were generally strongly positive between the diseases [for example, chr 2: 191051955 - 193033982].

**Fig. 5.**
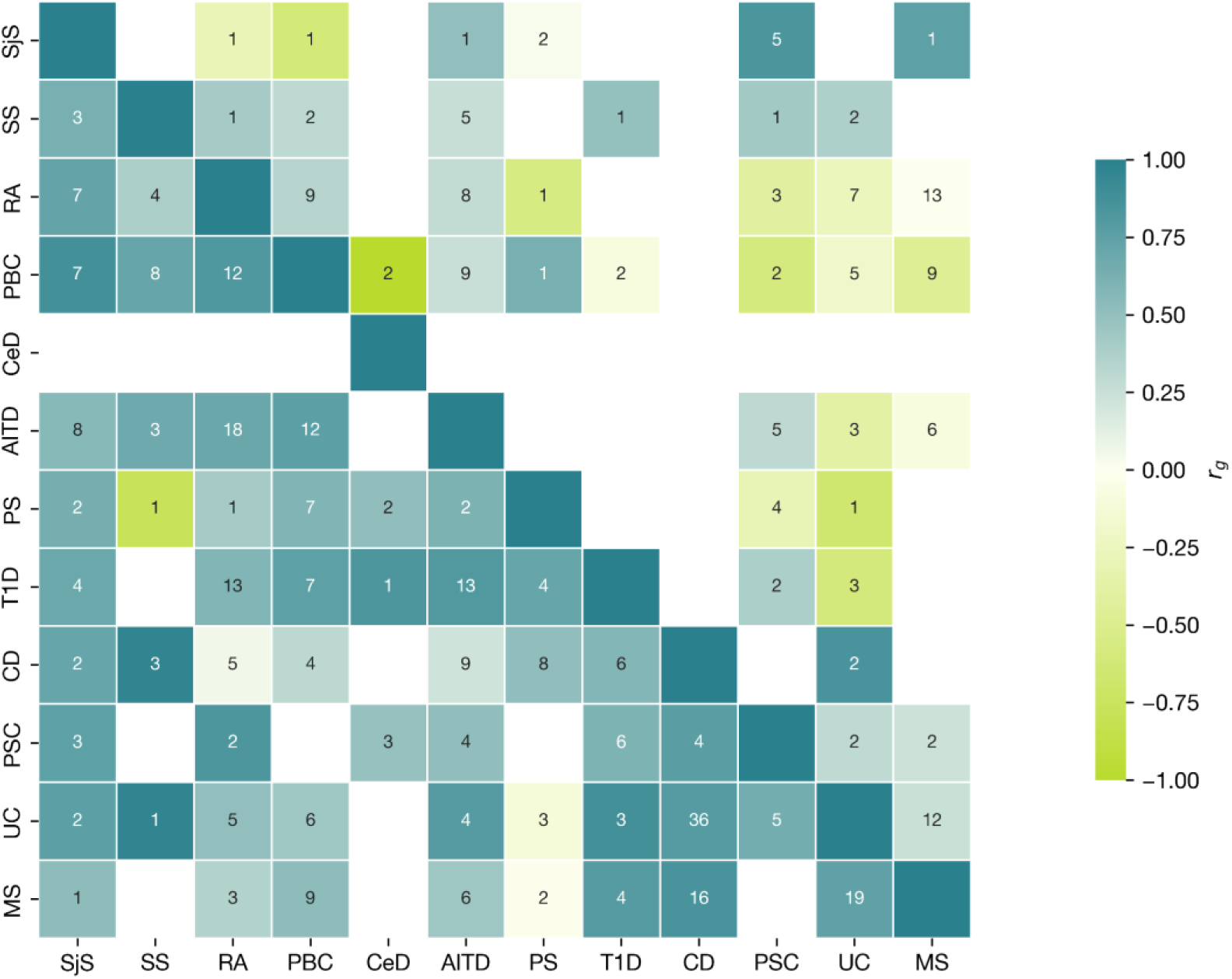
LAVA characteristics for MHC locus. Correlation matrix of positive and negative significant correlation, number of loci within MHC (upper triangular part), with loci outside MHC (bottom triangular part). Numbers in cells show the number of regions with significant heritability in both traits (Boferroni corrected), where the correlation was assessed. Colour indicates the direction of effect (Aquamarine - positive; green - negative) for mean across all correlation values (r_g_s) within the regions with significant h2 for each phenotype.

## 3.4. Functional Annotation

### 3.4.1. MAGMA shared gene results

MAGMA analysis for all 15 summary statistics revealed whole blood and spleen to be the most common tissues for all diseases except PSC, SjS and MG, and spleen for CeD. Also, Small Intestine Terminal Ileum, Cells_EBV-transformed_lymphocytes, and Lung participated in 11, 8 and 7 diseases respectively (Suppl. 2, Fig.5). We did not detect specific grouping of tissues by factor.

We assessed the most frequently associated genes across traits, which for 11 diseases were (Suppl. 2, Fig 4.): *GABBR1* (gamma-aminobutyric acid type B receptor subunit 1), *ZKSCAN3* (Zinc Finger With KRAB And SCAN Domains 3), PGBD1 (PiggyBac Transposable Element Derived 1) and for 10 diseases: *ZSCAN31* (Zinc Finger And SCAN Domain Containing 31), *SCAND3* (SCAN Domain Containing 3) and *ZSCAN23* (Zinc Finger And SCAN Domain Containing 23). We also revealed genes only included in factor 1 or factor 4 (the most margin groups of autoimmune and autoinflammatory phenotypes, Suppl. 1, Table 9). In factor 1, 22 genes were unique and in factor 4, 159 genes were unique with some of them involved in monogenic immune/autoinflammatory diseases (see Fig. 6. below, Suppl. 2., Fig 6. and Discussion section)

**Fig. 6.**
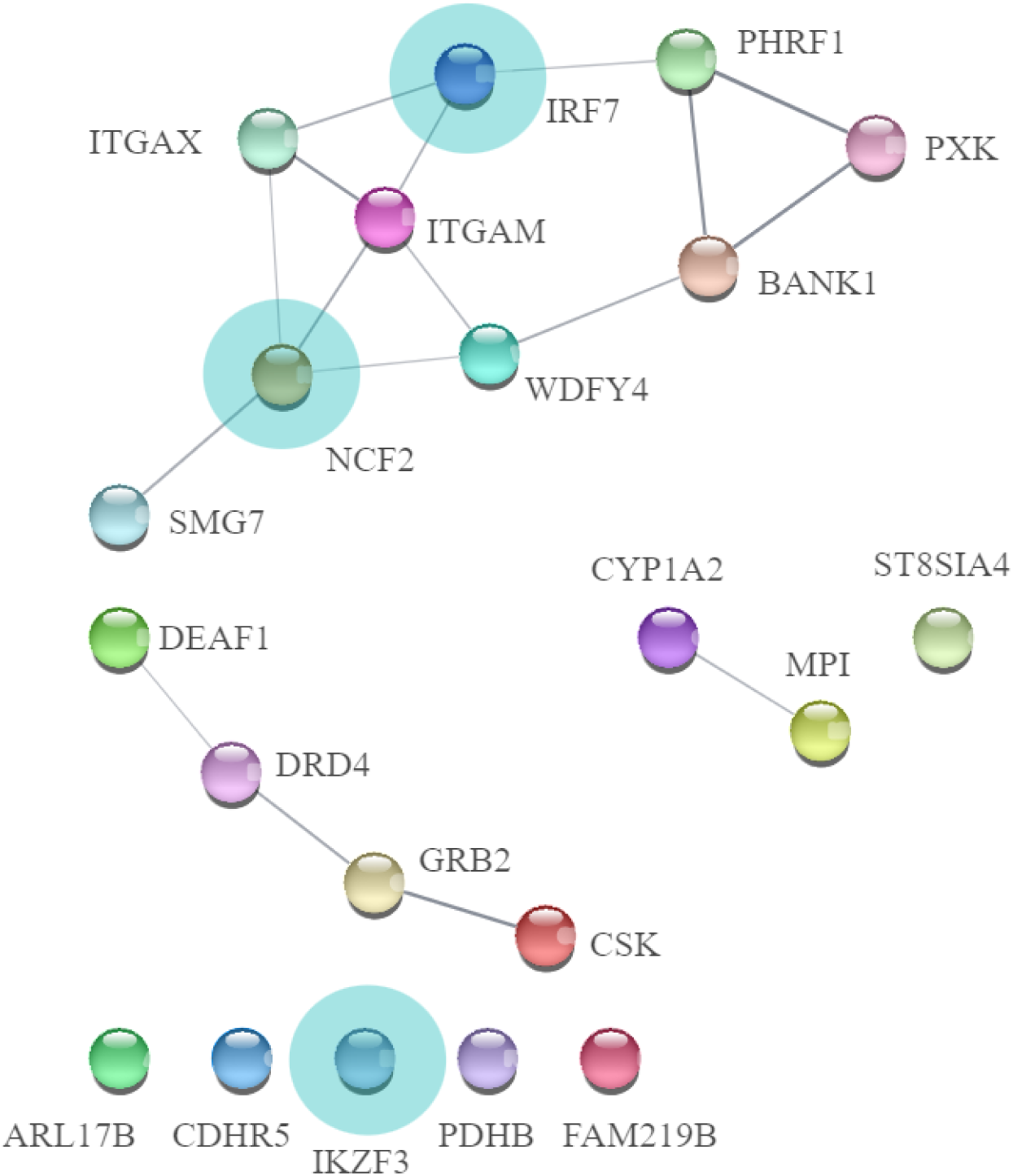
Genes uniquely involved in “autoimmune” factor 1. We marked genes linked to monogenic immune diseases with light blue circles. Lines between genes show the involvement in molecular networks (the stringApp for Cytoscape) Subsequently, we assessed gene sets and revealed the uniqueness of each factor. There were no gene sets specific for factor 2 because factor 2 diseases share common genes with other factors. For factor 1, 9 unique gene sets were revealed (N02-mediated IL12 pathways, PID_TCR_pathway, as well leukocytes and lymphocytes signalling), for factor 3 – 16 unique gene sets, for factor 4 – 41, and for out-models MS and T1D – 19 gene sets (Suppl. 1., Table 10 and discussion section).

## Discussion

In this study of the genetic architecture of diseases on the AIDs continuum, we observe a striking convergence of findings across different statistical genetics methods, which indicate clustering along the AIDs continuum. Here we present novel evidence to support the recent call [AutoCore network paper, Bousfiha et al., 2020, Guthrie et al., 2023 and TransImmunome project [Lorenzon et al., 2018, Tchitchek et al., 2024] for a paradigm of AIDs nosology to incorporate genetics of polygenic diseases.

We found that by using the state-of-the-art statistical approaches, it is possible to reveal four clusters across the AIDs continuum based on GWAS data, with diseases belonging to factor 1 (autoimmune) and diseases belonging to factor 4 (autoinflammatory). The extreme positions for factor 1 and factor 4 diseases were further supported by trivariate MiXeR analysis. Significant overlap was observed within factor 1 diseases (SS, SLE, and RA), while there was a smaller overlap between factor 1 diseases (SLE, SS) and the factor 4 diseases (UC).

The classification of certain diseases such as CD, UC, and PSC is consistent with a previous study by Demela et al., 2023, and remains in the same cluster. Demela’s study also utilises genomic SEM to demonstrate pathway convergence. While they suggest this factor corresponds to gastric disorders, we show that other gastroenterological phenotypes (PBC and CeD), cluster with different diseases (F1+3 and F3, respectively). Therefore, this grouping may not solely be due to involvement of the gastroenterological tract but may suggest grouping based on autoinflammatory pathogenesis. Topaloudi A. et al. [Topaloudi A. et al., 2023] also revealed four factors structure in line with a recent preprint [Breunig et al., 2024]. According to Topaloudi paper MS was in the same group as PSC and this connection in our paper was shown in LDSC and LAVA, as well as MG was in the same group as RA, and we were able to show that they share part of factor 2. Based on our findings and previous literature [Demela et al., 2023, Topaloudi et al. 2023, Williams et al., 2023] it appears that the factor structure in the genomic SEM model depends on 1) the initial set of disorders and 2) GWAS power. By including more diseases in our analyses compared to the previous studies, we were able to differentiate more factors within the autoimmune spectrum. This insight can enhance the classification of AIDs.

In the Transimmunome project [Tchitchek et al., 2024] five factors were revealed, but with inclusion of diseases with monogenic inheritance (Familial Mediterranean Fever) or diseases without powerful GWAS (idiopathic inflammatory myopathies). Additionally, our factor 3 consists of diseases which were not studied in their project. Tchitchek et al. revealed the belonging of CD and UC to the same factor which supports our findings on biomarkers level. According to Tchitchek’s classification, RA and T1D were in the same group, and we shown the significant overlap in MiXeR (rg 0.33, shared fraction 38%) and had strong local positive correlations in some loci as indicated by LAVA disregarding the fact that T1D is out of model in our study.

Also, in our study RA and SLE belong in factor one and RA in both factor 1 and 2, indicating that RA has a different genetic background compared to SLE. SLE is regarded as a classical autoimmune disease with autoantibody production, whereas RA has both autoimmune (classical seropositive RA) and mixed/autoinflammatory features (seronegative subtypes, Szekanecz et al., 2021). A notable observation is the grouping of MG together with CeD, which can help to reveal background for co-incidence of both diseases in case-reports without confirmed evidence at the population level [Thawani et al., 2018]. If we focus on MS, which were out of our genomic SEM model, but it was shown that MS has a high correlation and overlap with factor 4 diseases and PBC by other methods. That was previously shown [Kim et al., 2022] and support the fact that MS has common genetic background not only with autoimmune but also with autoinflammatory clusters.

LAVA analysis showed a mixed pattern (positive and negative) of correlations inside the MHC region in AIDs, but in loci outside MHC correlations were mostly positive. The same pattern for mixed correlation in the main MHC hotspot [chr6: 32539568 - 32586784] was shown for AIDs in the original LAVA paper [Werme et al., 2022]. Most MHC loci with significant local correlation belong to the MHC II subgroup. On the contrary PS which belongs to MHC-1-opathy according to EULAR classification [Kuiper et al., 2023, Matzaraki et al., 2017] in most of the cases has a negative correlation with other diseases in top-MHC loci. The MHC findings could be limited by non-specialized genotyping chips in GWAS and resolution of the MHC region, as well as limitations of the LAVA method due to LD structure and GWAS power.

The most frequent genes across diseases were involved in cell signalling and interaction and can be common for all types of immune disorders, consistent with Lincoln et al. (2024). Then we assessed the unique genes in marginal “autoimmune” and “autoinflammatory factors”. In “autoimmune” factor, 22 genes were unique, among them neutrophil cytosolic factor 2, which is associated with autosomal recessive chronic granulomatous disease 2 with neutrophils lacking superoxide-generating act [Nunoi et al., 1998]. Mutation in Interferon Regulatory Factor 7 was described as a cause of immunodeficiency-39 in a child [Ciancanelli, M. J. et al, 2015] and as a gene which is linked to intrinsic and innate immunity according to the classification of inborn errors of immunity [Tangye et al., 2022]. Mutation in IKZF3 was described in Immunodeficiency-84, which is an autosomal dominant primary immunologic disease associated with low levels of B cells and impaired early B-cell development [Yamashita, M. et al., 2021]. For “autoinflammatory” factor 4, 159 genes were unique, but less percentage participated in the immunological process [Suppl. 2., Fig. 6]. For example, the NOD2 gene plays a role in Blau syndrome, a rare autosomal dominant autoinflammatory syndrome classified as an autoinflammatory phenotype according to Tangye et al., 2022.These findings are in line with AutoCore’s idea that monogenic AIDs may represent genetically determined, more severe forms of more common polygenic AIDs. That also enables the transfer of knowledge between rare and complex diseases [Guthrie et al., 2023].

In the assessment of gene sets, we identified the involvement of Interleukin 12 (IL12) and IL23 gene sets in factor 4 diseases. A monoclonal antibody which targets IL12/IL23 (ustekinumab) is approved for the treatment of UC and CD as well as PS [Aggeletopoulou et al., 2018, Koutruba et al., 2010]. Also, IL3, IL4, IL18, IL27, T-helper and IFN gene sets involved in “autoinflammatory” factor 4 provide valuable insights that can inform the potential application of drugs that targets these interleukins and related pathways.

For factor 1, N02-mediated IL12 pathways in NK cells were involved as well as the IL12 pathway, and that was shown before to be associated with SLE genetic risk [Larosa M et al., 2019]. Also, TCR signalling in naïve CD4+ T cells and leukocyte-associated pathways participated in “autoimmune” factor 1. These pathways can also be important for SjS and SS where fewer studies about the involvement of IL pathways exist due to the lack of powerful data [Shi et al., 2024].

In factor 3 diseases a lot of pathways were involved linked to T-cells, B-cells and JAK pathways as well as IL17. Involvement of IL17 in JIA pathogenesis was recently shown in different studies [Paroli et al., 2022] as well secukinumab, Il17 blocker, was approved for treating of 2 of 7 JIA subtypes (juvenile psoriatic arthritis and enthesitis-related arthritis) as well as for PS treatment. Also, for MS and T1D which were out of any factors in genomic SEM, different IL2, IL6 and T-cell pathways were important.

The accumulated evidence from this study and previous findings can help inform immunotherapy for AIDs. Already designed drugs which suppress major pro-inflammatory signalling pathways as IL-17, JAK inhibitors have tremendous success compared to traditional systemic therapies. Unfortunately, there are still unaddressed medical requirements in terms of both long-term safety and overall effectiveness, as a considerable number of patients do not attain disease remission. Enhanced understanding of the genetically informed mechanisms and diversity within AIDs would create opportunities to address these challenges, resulting in a more personalised and efficient treatment approach.

A limitation of our study is that it only focussed on GWAS performed on populations of European ancestry. This is because genomic SEM and LD score regression require the samples to be drawn from the same ancestry, as well lack of powerful GWAS for non-European ancestry for most of the diseases. The second limitation was that we could use only powerful GWASes for polygenic disease for statistical methods available and had to ignore small GWASes as well as data freeze performed before analysis (August 2023). Also, cohort sample size and SNP-sets are drivers of the limited resolution of genetic mapping and the ability to detect robust disease associations [Uffelmann et al. 2021]. An additional limitation was to focus only on genetics but not on the deep immunophenotyping via cells/immune molecules due to the lack of powerful data for these phenotypes. Our findings about genetic clusters and overlap does not necessarily disqualify the current clinical system but bring attention to distribution of genetic factors. As well data on infections, vaccine regimen and the treatment with anti-inflammatory drugs lack in used GWAS and that is a common limitation of studies performed on summary statistics data.

To summarise, the revealing of four-factors-clustering can help to define the disease groups for which it is possible to use the same therapies. Furthermore, our results can be instrumental in assembling cluster-guided multi-parametric analyses that include genetics and omics data and enable deep phenotyping of patients leading to personalised drug selection. The current study sheds light on the autoimmune-autoinflammatory continuum from a genetic perspective and can inform future studies in this field.

## Data Availability

GWAS data (summary statistics) are publicly available on GWAS catalog https://www.ebi.ac.uk/gwas/ or FinnGen database https://www.finngen.fi/en, or IMSGC database https://imsgc.net/.
All data produced in the present study are available upon reasonable request to the authors.

https://www.finngen.fi/en

https://imsgc.net/

## Acknowledgements

All GWAS investigated in the present study were approved by the local ethics committees, and informed consent was obtained from all participants. The authors thank the researchers of the IMSGC https://imsgc.net/, and consortia for access to data, and for all participants who provided DNA samples. We want to acknowledge the participants and investigators of the FinnGen study: https://www.finngen.fi/ [Kurki MI et al., 2023].

We gratefully acknowledge support from the the Research Council of Norway (#223273, 248778, 300309, 324252, 324499, 326813, 344121), European Union’s Horizon 2020 research and innovation programme under the Marie Skłodowska-Curie Actions Grant 801133 (Scientia fellowship); the South-East Norway Regional Health Authority (2022-073 and 2023-087), KG Jebsen Stiftelsen. This project has received funding from the European Union’s Horizon 2020 research and innovation programme under grant agreement No 847776 and 964874, and EEA grants (EEA-RO-NO-2018-0535, EEA-RO-NO-2018-0573).

This work was partly performed on the TSD (Tjeneste for Sensitive Data) facilities, owned by the University of Oslo and operated and developed by the TSD service group at the University of Oslo, IT Department (USIT), with resources provided by UNINETT Sigma2 - the National Infrastructure for High Performance Computing and Data Storage in Norway.

Computations were also performed on resources provided by the National Infrastructure for High-Performance Computing and Data Storage in Norway.

## Data availability and Computational tools

GWAS data are publicly available or available on request.

Statistical analyses were performed with Python and R using existing tools available on FUMA https://fuma.ctglab.nl/

FUMA https://fuma.ctglab.nl/

GitHub https://github.com/precimed/

Genomic SEM https://github.com/GenomicSEM/

METAL https://github.com/statgen/METAL

MiXeR bivariate https://github.com/precimed/mixer

MiXeR trivariate https://codeberg.org/intercm/mix3r

LAVA https://github.com/josefin-werme/LAVA

LDSC https://github.com/bulik/ldsc

MAGMA https://cncr.nl/research/magma/

Python Convert https://github.com/precimed/python_convert

## Abbreviations

AD: autoimmune diseases
AIC: Akaike Information Criterion
AIDs: autoimmune and autoinflammatory disorders
AIF: autoinflammatory disorders
AITD: autoimmune thyroiditis
EFA: exploratory and confirmatory factor analysis
CeD: celiac disease
CD: Crohn’s disease
CFA: confirmatory factor analysis
CFI: Comparative Fit Index
CNS: central nervous system
Genomic SEM: genomic structural equation modelling
IL: interleukin
JAK: Janus kinase
JIA: juvenile idiopathic arthritis
LAVA: Local Analysis of [co]Variant Association
LD: linkage disequilibrium
LDSC: linkage disequilibrium score regression
MHC: major histocompatibility complex
MS: multiple sclerosis
MiXeR: Gaussian causal mixture modelling
MG: myasthenia gravis
PBC: primary biliary cholangitis
PS: psoriasis
PSC: primary sclerosing cholangitis
RA: rheumatoid arthritis
SjS: primary Sjogren’s syndrome
SLE: systemic lupus erythematosus
SRMR: standardised root mean square residual
SS: systemic sclerosis
T1D: type 1 diabetes
UC: ulcerative colitis

