## Supplementary_2 for "Mapping the genetic landscape of disorders on the autoimmune-autoinflammatory continuum: potential implications for classification and treatment"

**Supplementary 2., Fig. 1. Heatmap of genome-wide genetic correlations across 15 immune-mediated diseases.** A. Numbers in the cells show genetic correlations estimated using LD score regression method. Blue and red indicate positive and negative correlations respectively. B. Green and purple indicate positive and negative correlations respectively, asterisk means significant correlation between phenotypes (Bonferroni correction to number of comparisons).

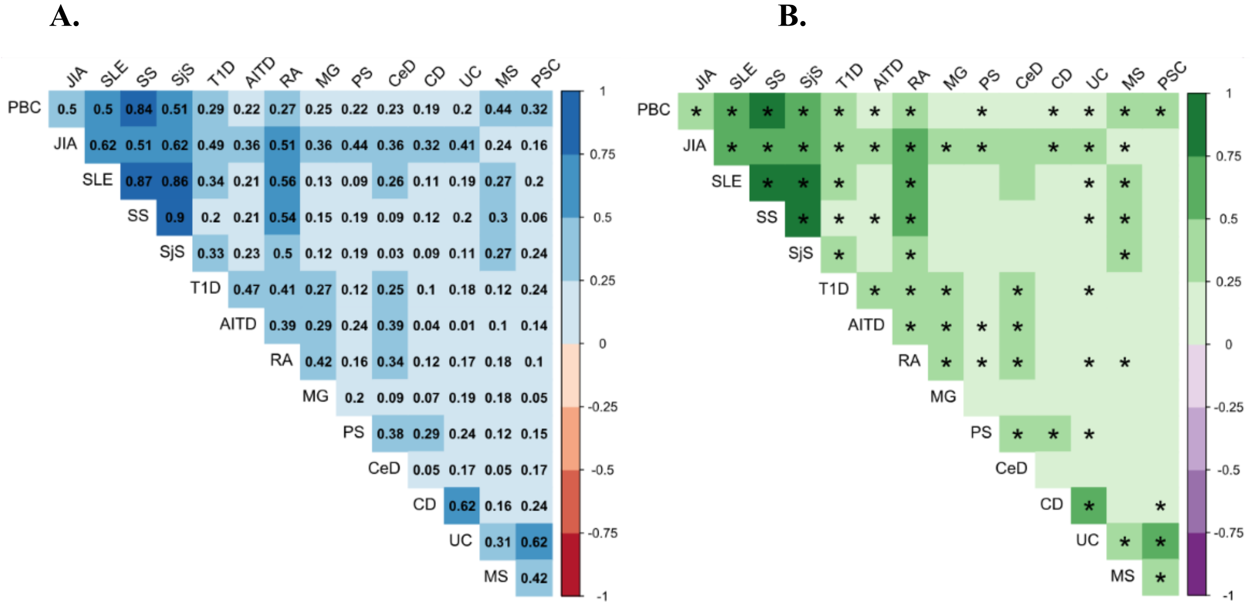


**Supplementary 2., Fig. 2. Genetic overlap beyond genetic correlations**.

Fraction of disease-influencing genetic variants shared between two diseases with respect to the less polygenic disease in the pair estimated with Bivariate Gaussian causal mixture model (MiXeR) for 15 immune-mediated diseases.

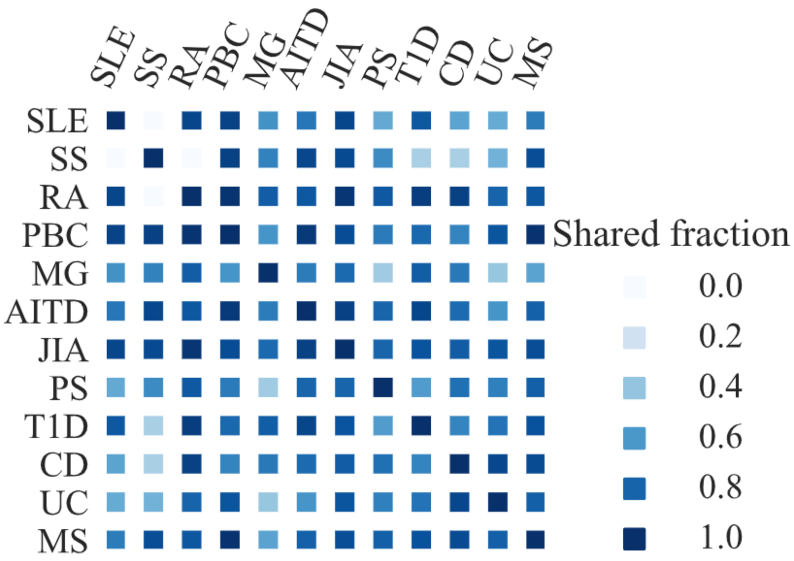

**Supplementary 2., Fig. 3. Local genetic correlations across immune-mediated diseases.** Heatmap of local genetic correlations estimated with LAVA. The color indicates average correlation across all positively (top right) and negatively (bottom left) correlated regions. Numbers in cells indicate the number of regions with significant local generic correlation. The absence of number indicates that there were no regions with significant correlation.

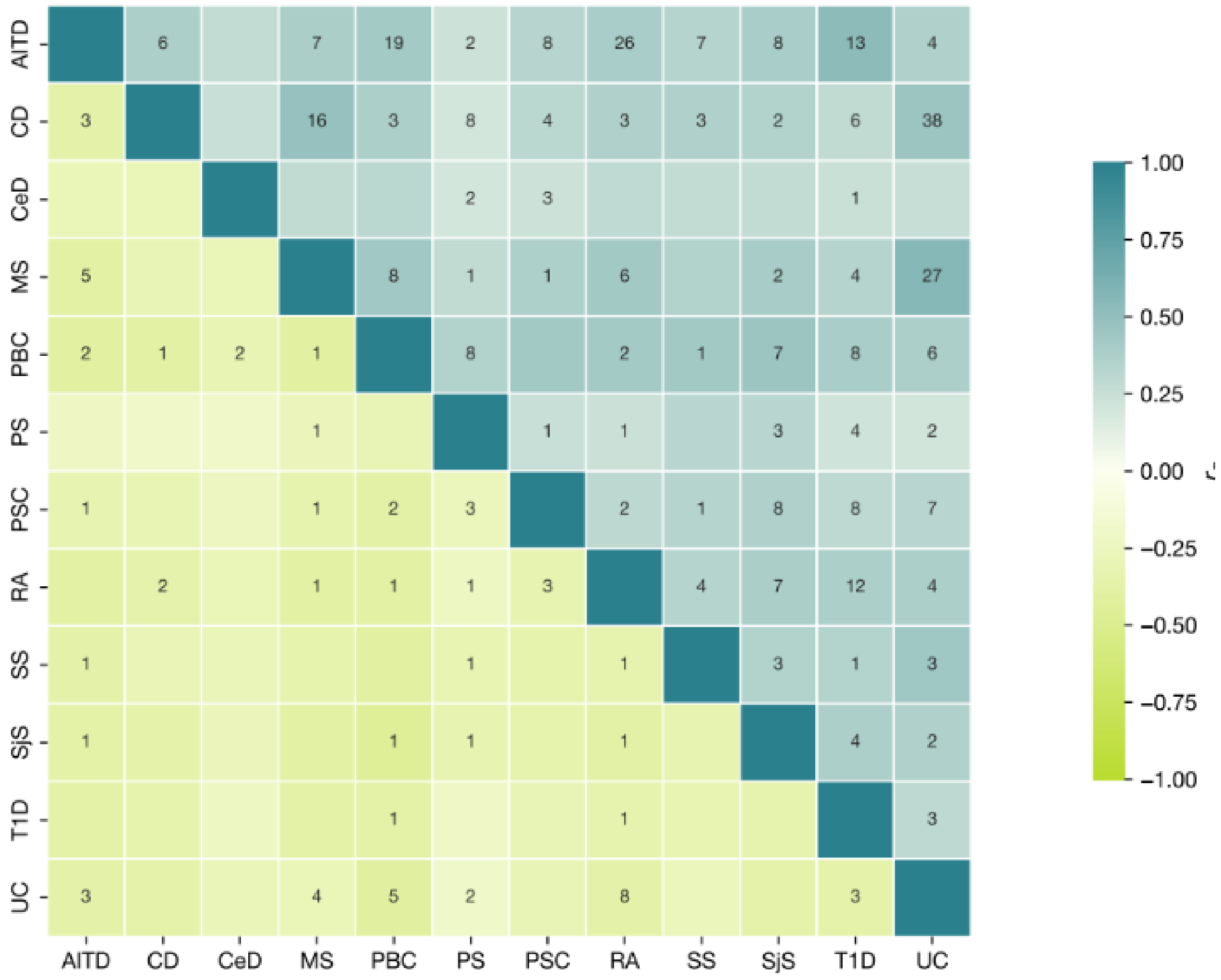

**Supplementary 2., Fig. 4. Genes shared between five or more immune-mediated diseases.** For each disease, associated genes were identified using MAGMA.

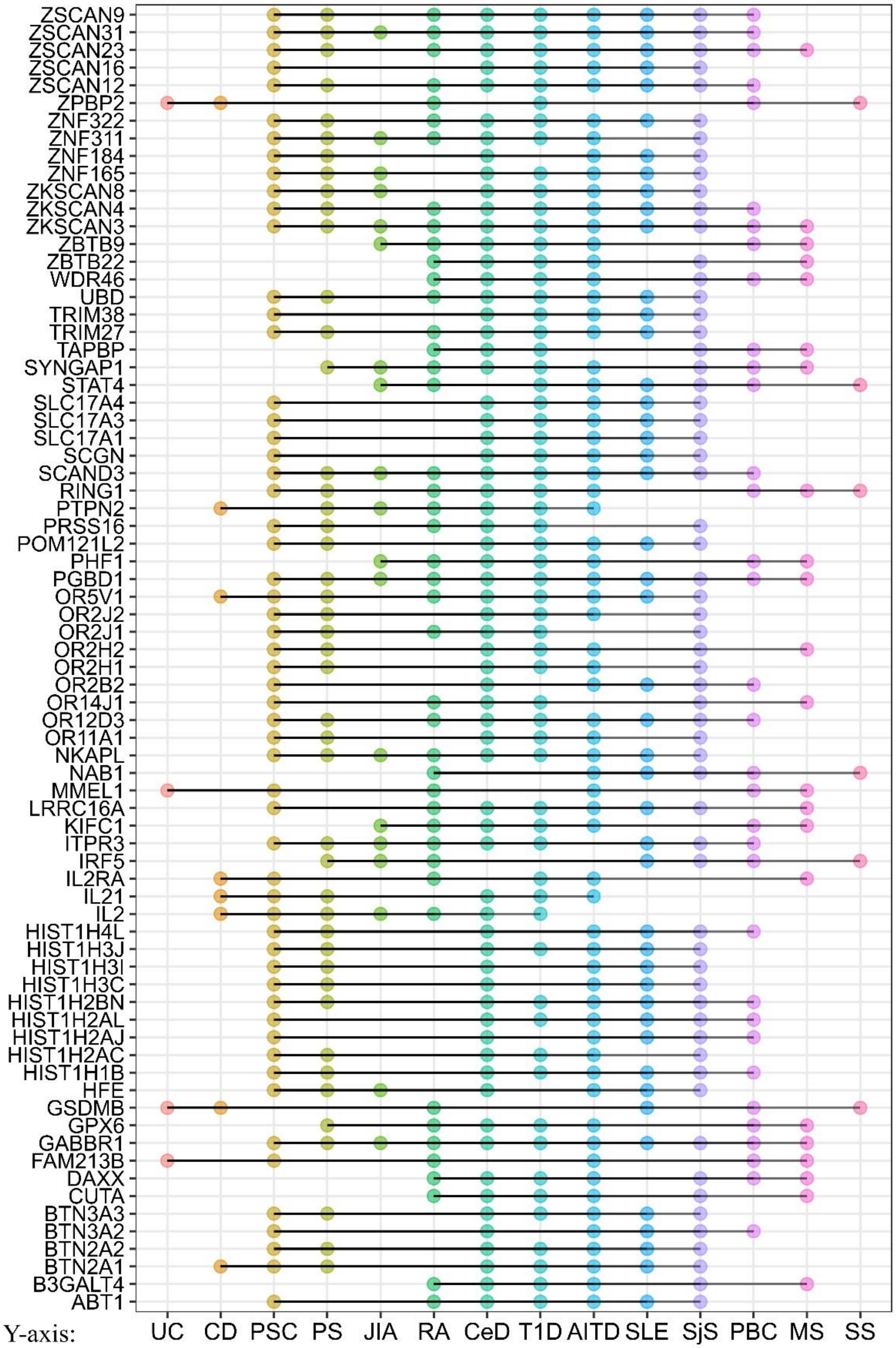


Y-axis:

Gene names

**Supplementary 2., Fig. 5. Tissue specificity analysis.** For each disease, genes identified in MAGMA analysis were tested for tissue specific expression using gene expression data from 54 tissue types available in GTEx v8. Significant results are highlighted in red color. Analysis was performed using FUMA GENE2FUNC.

**SjS**

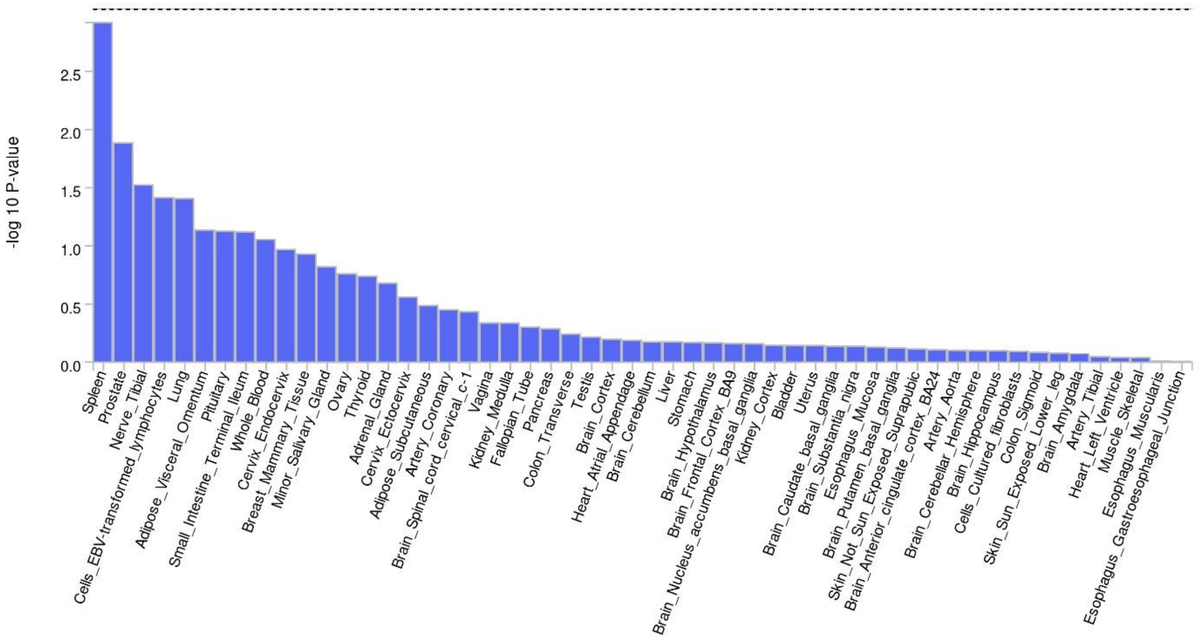


**SLE**

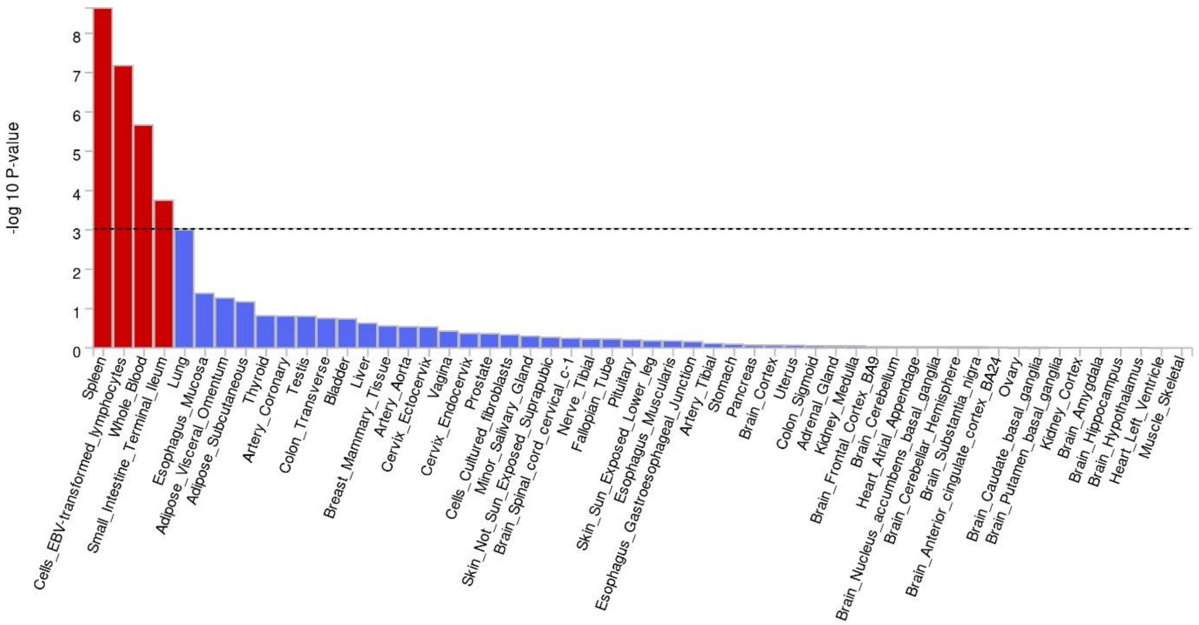

**SS**

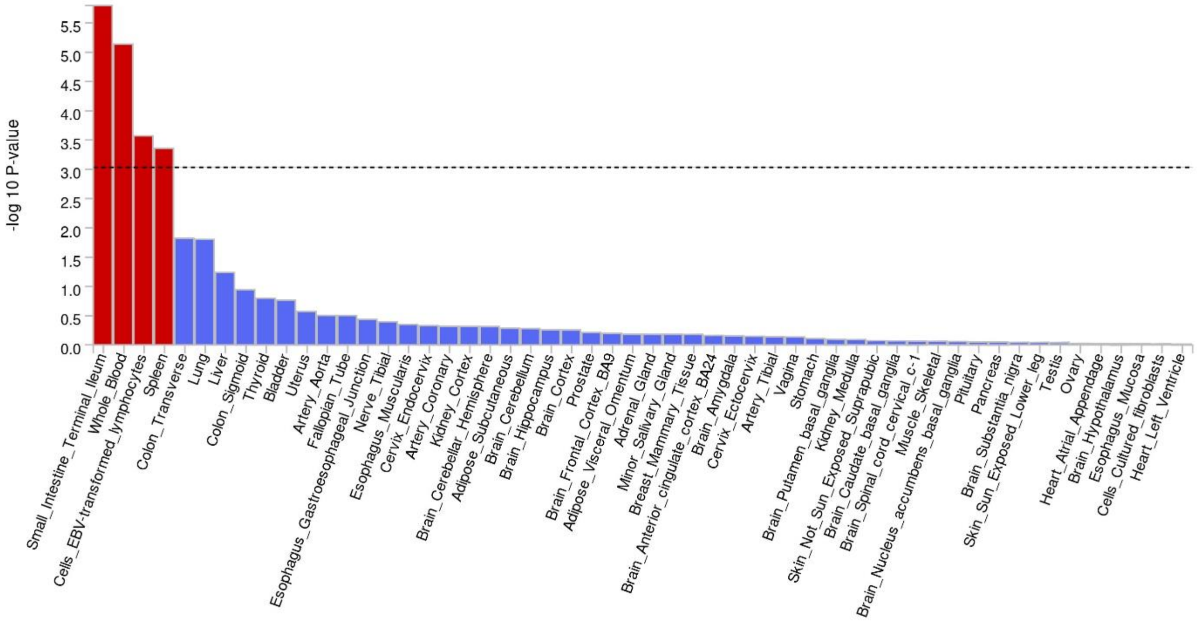

-log 10 P-value
5.5
5.0
4.5
4.0
3.5
3.0
2.5
2.0
1.5
1.0
0.5
0.0
Small_Intestine_Terminal_Ileum
Whole_Blood
Cells_EBV-transformed_lymphocytes
Spleen
Colon_Transverse
Lung
Liver
Colon_Sigmoid
Thyroid
Bladder
Uterus
Artery_Aorta
Fallopian_Tube
Esophagus_Gastroesophageal_Junction
Nerve_Tibial
Esophagus_Muscularis
Cervix_Endocervix
Artery_Coronary
Kidney_Cortex
Brain_Cerebellar_Hemisphere
Adipose_Subcutaneous
Brain_Cerebellum
Brain_Hippocampus
Brain_Cortex
Prostate
Brain_Frontal_Cortex_BA9
Adipose_Visceral_Omentum
Adrenal_Gland
Minor_Salivary_Gland
Breast_Mammary_Tissue
Brain_Anterior_cingulate_cortex_BA24
Brain_Amygdala
Cervix_Ectocervix
Artery_Tibial
Vagina
Stomach
Brain_Putamen_basal_ganglia
Kidney_Medulla
Skin_Not_Sun_Exposed_Suprapubic
Brain_Caudate_basal_ganglia
Brain_Spinal_cord_cervical_c-1
Muscle_Skeletal
Brain_Nucleus_accumbens_basal_ganglia
Pituitary
Pancreas
Brain_Substantia_nigra
Skin_Sun_Exposed_Lower_leg
Testis
Ovary
Heart_Atrial_Appendage
Brain_Hypothalamus
Esophagus_Mucosa
Cells_Cultured_fibroblasts
Heart_Left_Ventricle


**PBC**

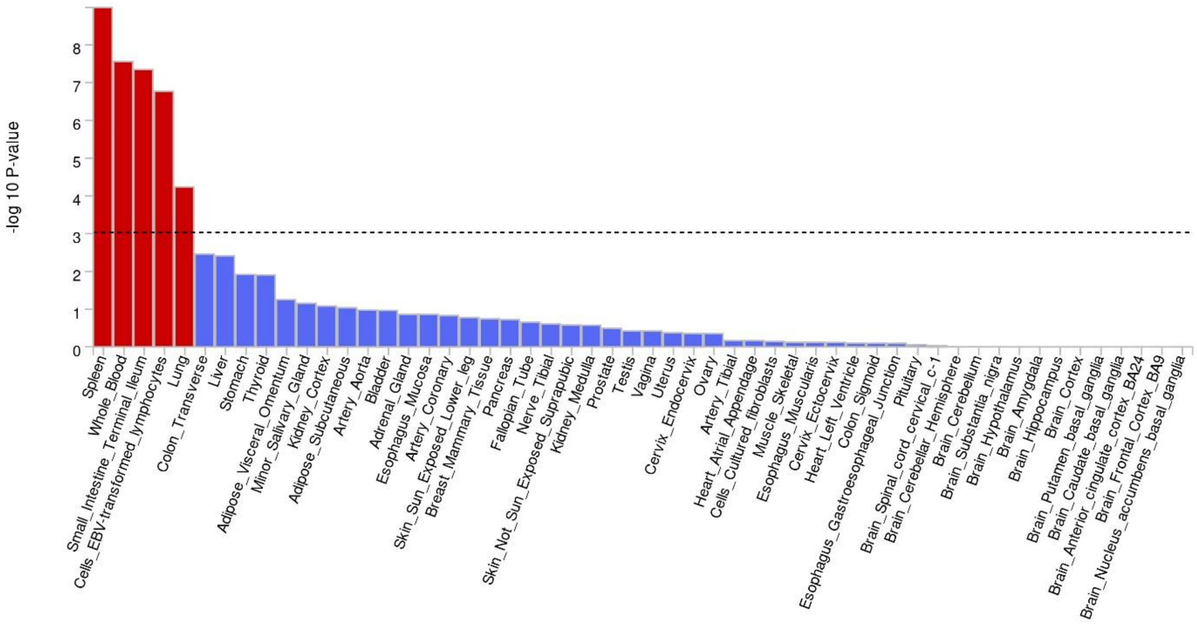

-log 10 P-value
8
7
6
5
4
3
2
1
0
Spleen
Whole_Blood
Small_Intestine_Terminal_Ileum
Cells_EBV-transformed_lymphocytes
Lung
Colon_Transverse
Liver
Stomach
Thyroid
Adipose_Visceral_Omentum
Minor_Salivary_Gland
Kidney_Cortex
Adipose_Subcutaneous
Artery_Aorta
Bladder
Adrenal_Gland
Esophagus_Mucosa
Artery_Coronary
Skin_Sun_Exposed_Lower_leg
Breast_Mammary_Tissue
Pancreas
Fallopian_Tube
Nerve_Tibial
Skin_Not_Sun_Exposed_Suprapubic
Kidney_Medulla
Prostate
Testis
Vagina
Uterus
Cervix_Endocervix
Ovary
Artery_Tibial
Heart_Atrial_Appendage
Cells_Cultured_fibroblasts
Muscle_Skeletal
Esophagus_Muscularis
Cervix_Ectocervix
Heart_Left_Ventricle
Colon_Sigmoid
Esophagus_Gastroesophageal_Junction
Pituitary
Brain_Spinal_cord_cervical_c-1
Brain_Cerebellar_Hemisphere
Brain_Cerebellum
Brain_Substantia_nigra
Brain_Hypothalamus
Brain_Amygdala
Brain_Hippocampus
Brain_Cortex
Brain_Putamen_basal_ganglia
Brain_Caudate_basal_ganglia
Brain_Anterior_cingulate_cortex_BA24
Brain_Frontal_Cortex_BA9
Brain_Nucleus_accumbens_basal_ganglia

**CeD**

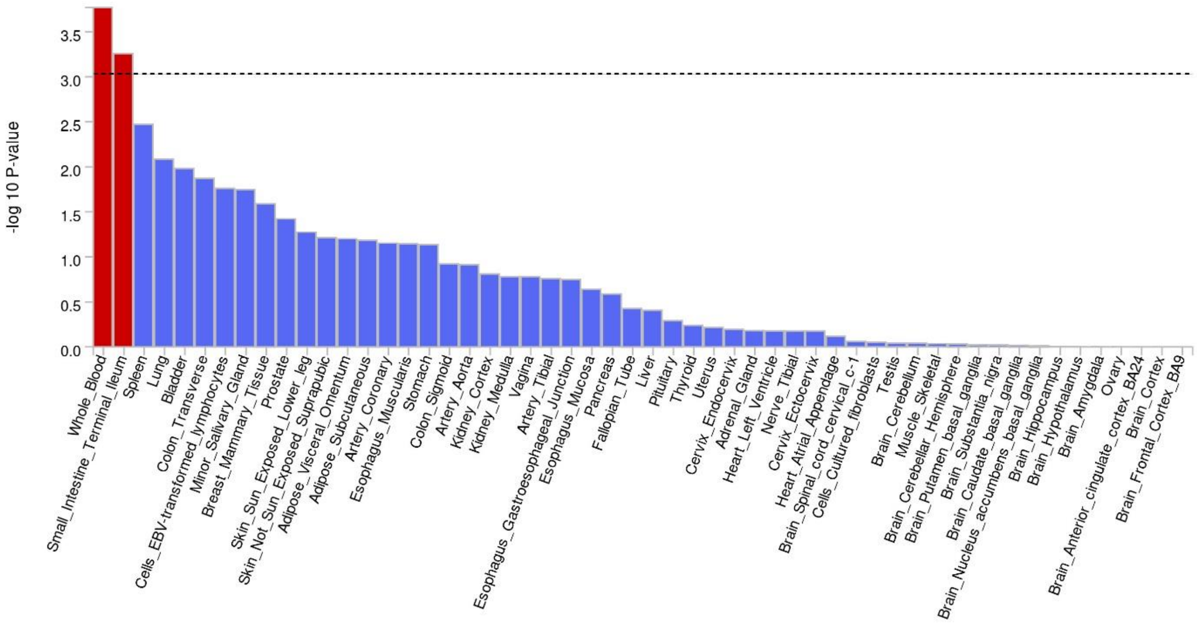
-log 10 P-value
Whole_Blood
Small_Intestine_Terminal_Ileum
Spleen
Lung
Bladder
Colon_Transverse
Cells_EBV-transformed_lymphocytes
Minor_Salivary_Gland
Breast_Mammary_Tissue
Prostate
Skin_Sun_Exposed_Lower_leg
Skin_Not_Sun_Exposed_Suprapubic
Adipose_Visceral_Omentum
Adipose_Subcutaneous
Artery_Coronary
Esophagus_Muscularis
Stomach
Colon_Sigmoid
Artery_Aorta
Kidney_Cortex
Kidney_Medulla
Vagina
Artery_Tibial
Esophagus_Gastroesophageal_Junction
Esophagus_Mucosa
Pancreas
Fallopian_Tube
Liver
Pituitary
Thyroid
Uterus
Cervix_Endocervix
Adrenal_Gland
Heart_Left_Ventricle
Nerve_Tibial
Cervix_Ectocervix
Heart_Atrial_Appendage
Brain_Spinal_cord_cervical_c-1
Cells_Cultured_fibroblasts
Testis
Brain_Cerebellum
Muscle_Skeletal
Brain_Cerebellar_Hemisphere
Brain_Putamen_basal_ganglia
Brain_Substantia_nigra
Brain_Caudate_basal_ganglia
Brain_Nucleus_accumbens_basal_ganglia
Brain_Hippocampus
Brain_Hypothalamus
Brain_Amygdala
Ovary
Brain_Anterior_cingulate_cortex_BA24
Brain_Cortex
Brain_Frontal_Cortex_BA9

**RA**

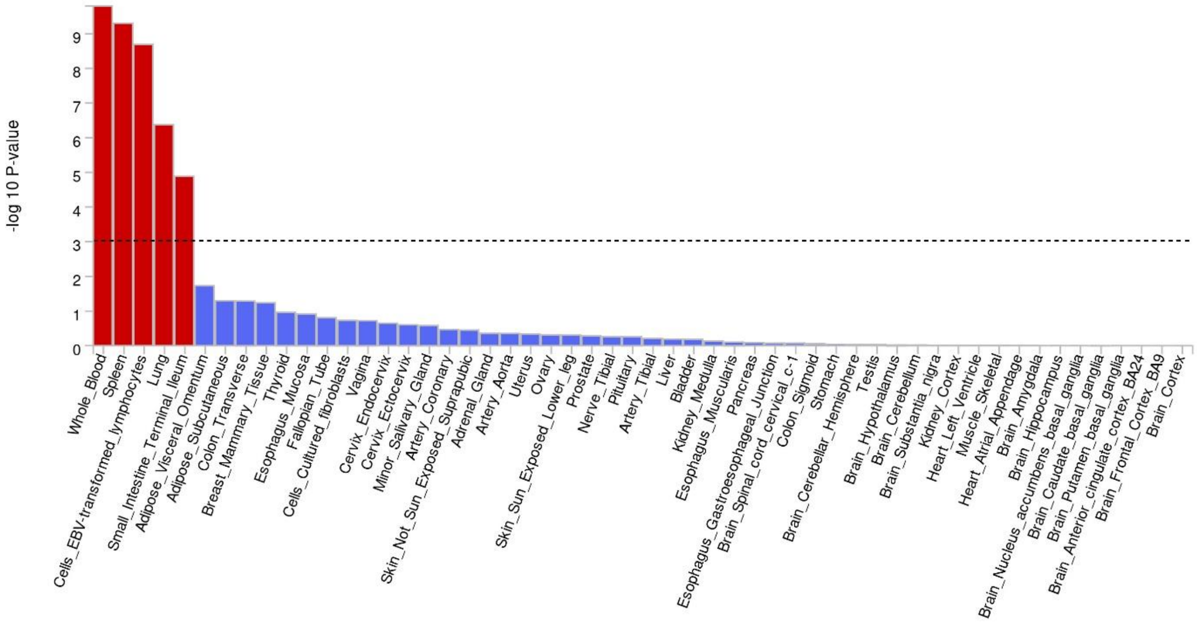
-log 10 P-value
Whole_Blood
Spleen
Cells_EBV-transformed_lymphocytes
Lung
Small_Intestine_Terminal_Ileum
Adipose_Visceral_Omentum
Adipose_Subcutaneous
Colon_Transverse
Breast_Mammary_Tissue
Thyroid
Esophagus_Mucosa
Fallopian_Tube
Cells_Cultured_fibroblasts
Vagina
Cervix_Endocervix
Cervix_Ectocervix
Minor_Salivary_Gland
Artery_Coronary
Skin_Not_Sun_Exposed_Suprapubic
Adrenal_Gland
Artery_Aorta
Uterus
Ovary
Skin_Sun_Exposed_Lower_leg
Prostate
Nerve_Tibial
Pituitary
Artery_Tibial
Liver
Bladder
Kidney_Medulla
Esophagus_Muscularis
Pancreas
Esophagus_Gastroesophageal_Junction
Brain_Spinal_cord_cervical_c-1
Colon_Sigmoid
Stomach
Brain_Cerebellar_Hemisphere
Testis
Brain_Hypothalamus
Brain_Cerebellum
Brain_Substantia_nigra
Kidney_Cortex
Heart_Left_Ventricle
Muscle_Skeletal
Heart_Atrial_Appendage
Brain_Amygdala
Brain_Hippocampus
Brain_Nucleus_accumbens_basal_ganglia
Brain_Caudate_basal_ganglia
Brain_Putamen_basal_ganglia
Brain_Anterior_cingulate_cortex_BA24
Brain_Frontal_Cortex_BA9
Brain_Cortex

**MG**

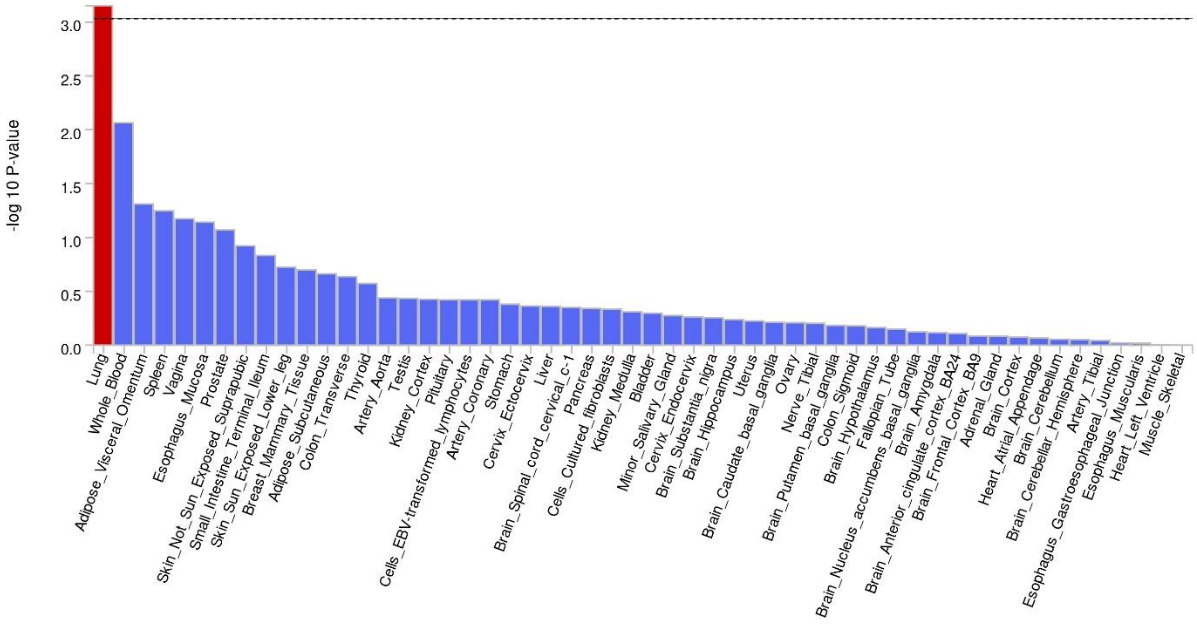
-log 10 P-value
0.0
0.5
1.0
1.5
2.0
2.5
3.0
Lung
Whole_Blood
Adipose_Visceral_Omentum
Spleen
Vagina
Esophagus_Mucosa
Prostate
Skin_Not_Sun_Exposed_Suprapubic
Small_Intestine_Terminal_Ileum
Skin_Sun_Exposed_Lower_leg
Breast_Mammary_Tissue
Adipose_Subcutaneous
Colon_Transverse
Thyroid
Artery_Aorta
Testis
Kidney_Cortex
Pituitary
Cells_EBV-transformed_lymphocytes
Artery_Coronary
Stomach
Cervix_Ectocervix
Liver
Brain_Spinal_cord_cervical_c-1
Pancreas
Cells_Cultured_fibroblasts
Kidney_Medulla
Bladder
Minor_Salivary_Gland
Cervix_Endocervix
Brain_Substantia_nigra
Brain_Hippocampus
Uterus
Brain_Caudate_basal_ganglia
Ovary
Nerve_Tibial
Brain_Putamen_basal_ganglia
Colon_Sigmoid
Brain_Hypothalamus
Fallopian_Tube
Brain_Nucleus_accumbens_basal_ganglia
Brain_Amygdala
Brain_Anterior_cingulate_cortex_BA24
Brain_Frontal_Cortex_BA9
Adrenal_Gland
Brain_Cortex
Heart_Atrial_Appendage
Brain_Cerebellum
Brain_Cerebellar_Hemisphere
Artery_Tibial
Esophagus_Gastroesophageal_Junction
Esophagus_Muscularis
Heart_Left_Ventricle
Muscle_Skeletal

**AITD**

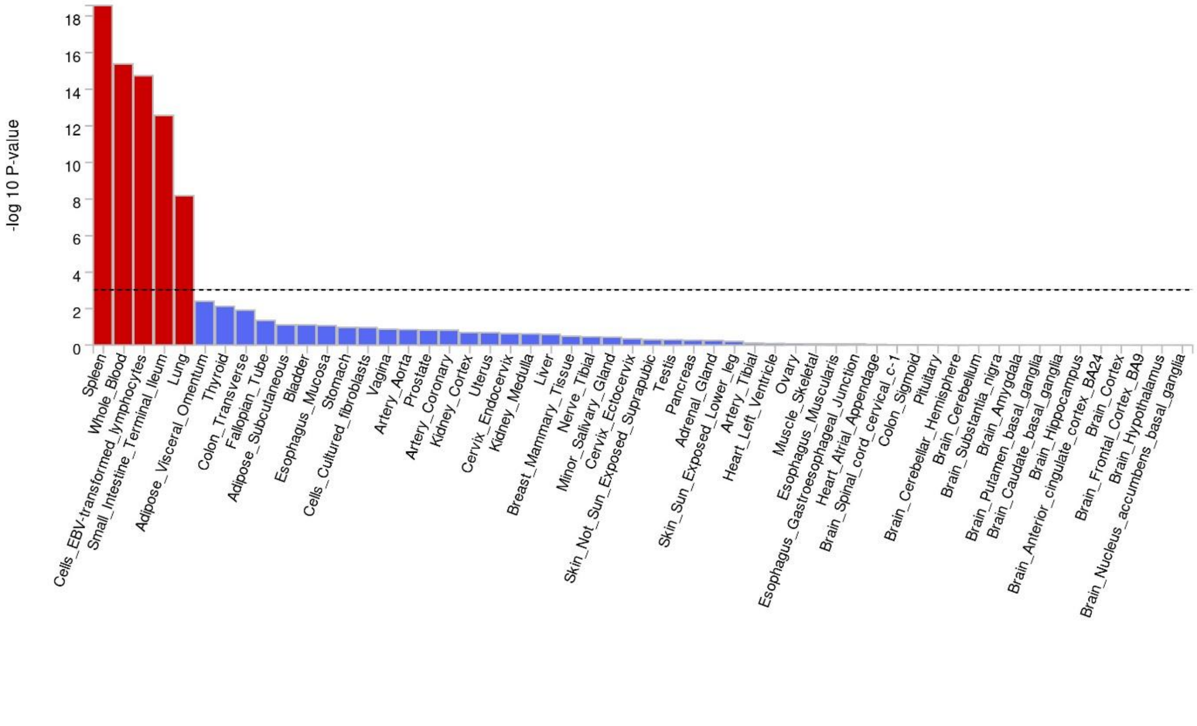
-log 10 P-value
0
2
4
6
8
10
12
14
16
18
Spleen
Whole_Blood
Cells_EBV-transformed_lymphocytes
Small_Intestine_Terminal_Ileum
Lung
Adipose_Visceral_Omentum
Thyroid
Colon_Transverse
Fallopian_Tube
Adipose_Subcutaneous
Bladder
Esophagus_Mucosa
Stomach
Cells_Cultured_fibroblasts
Vagina
Artery_Aorta
Prostate
Artery_Coronary
Kidney_Cortex
Uterus
Cervix_Endocervix
Kidney_Medulla
Liver
Breast_Mammary_Tissue
Nerve_Tibial
Minor_Salivary_Gland
Cervix_Ectocervix
Skin_Not_Sun_Exposed_Suprapubic
Testis
Pancreas
Adrenal_Gland
Skin_Sun_Exposed_Lower_leg
Artery_Tibial
Heart_Left_Ventricle
Ovary
Muscle_Skeletal
Esophagus_Muscularis
Esophagus_Gastroesophageal_Junction
Heart_Atrial_Appendage
Brain_Spinal_cord_cervical_c-1
Colon_Sigmoid
Pituitary
Brain_Cerebellar_Hemisphere
Brain_Cerebellum
Brain_Substantia_nigra
Brain_Amygdala
Brain_Putamen_basal_ganglia
Brain_Caudate_basal_ganglia
Brain_Hippocampus
Brain_Anterior_cingulate_cortex_BA24
Brain_Cortex
Brain_Frontal_Cortex_BA9
Brain_Hypothalamus
Brain_Nucleus_accumbens_basal_ganglia

**JIA**

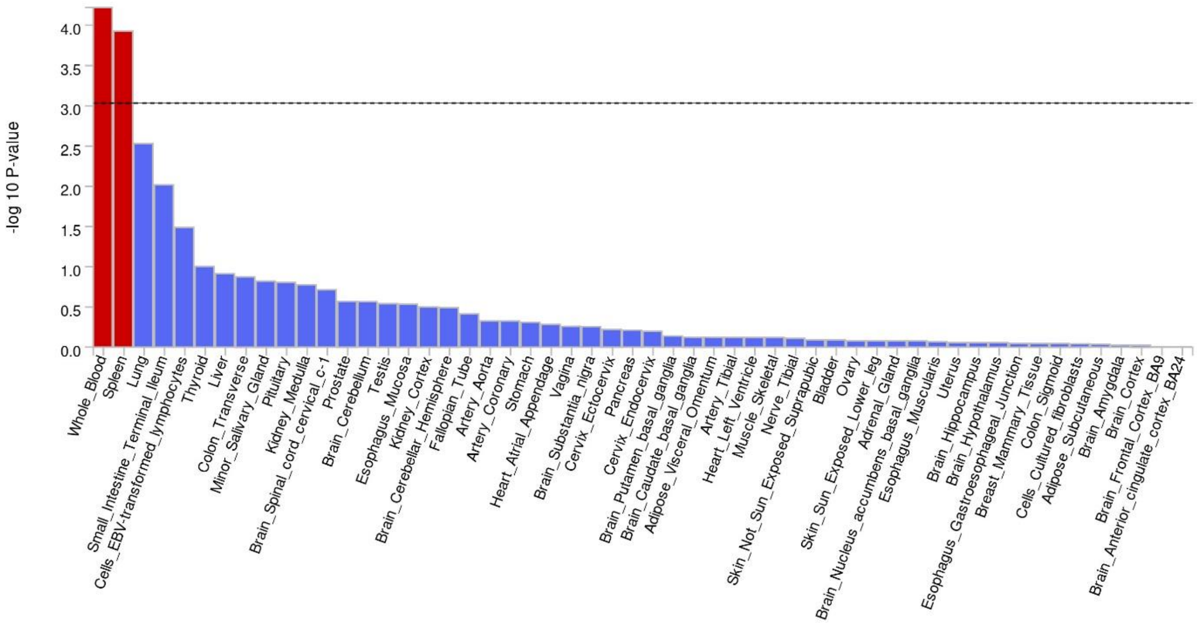
-log 10 P-value
Whole_Blood
Spleen
Lung
Small_Intestine_Terminal_Ileum
Cells_EBV-transformed_lymphocytes
Thyroid
Liver
Colon_Transverse
Minor_Salivary_Gland
Pituitary
Kidney_Medulla
Brain_Spinal_cord_cervical_c-1
Prostate
Brain_Cerebellum
Testis
Esophagus_Mucosa
Kidney_Cortex
Brain_Cerebellar_Hemisphere
Fallopian_Tube
Artery_Aorta
Artery_Coronary
Stomach
Heart_Atrial_Appendage
Vagina
Brain_Substantia_nigra
Cervix_Ectocervix
Pancreas
Cervix_Endocervix
Brain_Putamen_basal_ganglia
Brain_Caudate_basal_ganglia
Adipose_Visceral_Omentum
Artery_Tibial
Heart_Left_Ventricle
Muscle_Skeletal
Nerve_Tibial
Skin_Not_Sun_Exposed_Suprapubic
Bladder
Ovary
Skin_Sun_Exposed_Lower_leg
Adrenal_Gland
Brain_Nucleus_accumbens_basal_ganglia
Esophagus_Muscularis
Uterus
Brain_Hippocampus
Brain_Hypothalamus
Esophagus_Gastroesophageal_Junction
Breast_Mammary_Tissue
Colon_Sigmoid
Cells_Cultured_fibroblasts
Adipose_Subcutaneous
Brain_Amygdala
Brain_Cortex
Brain_Frontal_Cortex_BA9
Brain_Anterior_cingulate_cortex_BA24

**PS**

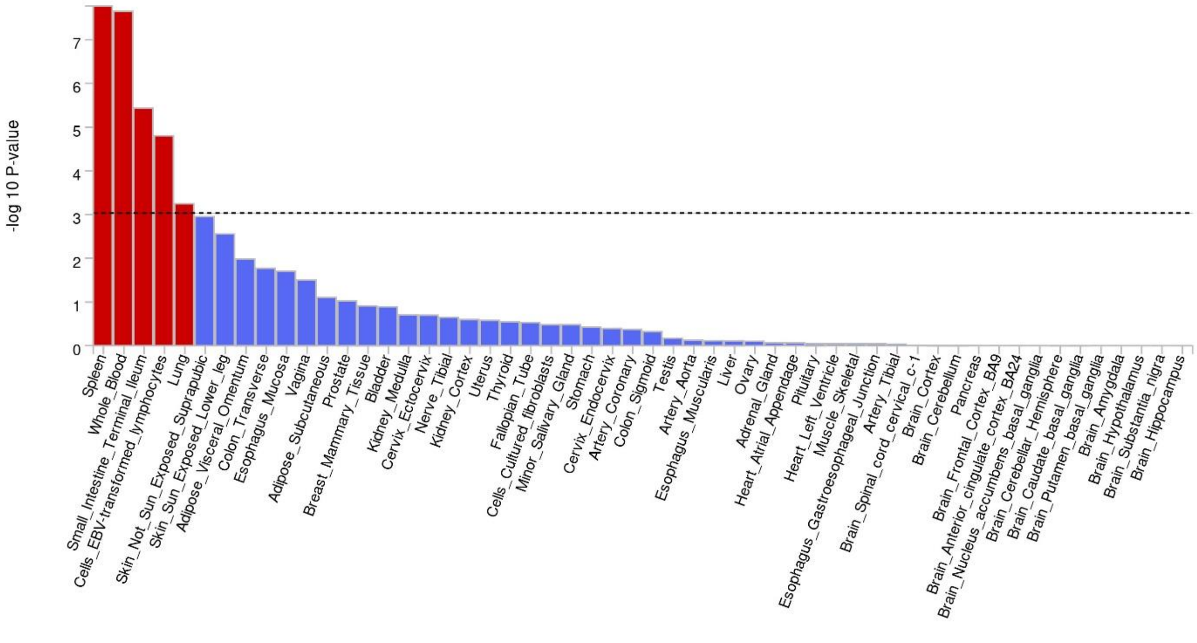
-log 10 P-value
Spleen
Whole_Blood
Small_Intestine_Terminal_Ileum
Cells_EBV-transformed_lymphocytes
Lung
Skin_Not_Sun_Exposed_Suprapubic
Skin_Sun_Exposed_Lower_leg
Adipose_Visceral_Omentum
Colon_Transverse
Esophagus_Mucosa
Vagina
Adipose_Subcutaneous
Prostate
Breast_Mammary_Tissue
Bladder
Kidney_Medulla
Cervix_Ectocervix
Nerve_Tibial
Kidney_Cortex
Uterus
Thyroid
Fallopian_Tube
Cells_Cultured_fibroblasts
Minor_Salivary_Gland
Stomach
Cervix_Endocervix
Artery_Coronary
Colon_Sigmoid
Testis
Artery_Aorta
Esophagus_Muscularis
Liver
Ovary
Adrenal_Gland
Heart_Atrial_Appendage
Pituitary
Heart_Left_Ventricle
Muscle_Skeletal
Esophagus_Gastroesophageal_Junction
Artery_Tibial
Brain_Spinal_cord_cervical_c-1
Brain_Cortex
Brain_Cerebellum
Pancreas
Brain_Frontal_Cortex_BA9
Brain_Anterior_cingulate_cortex_BA24
Brain_Nucleus_accumbens_basal_ganglia
Brain_Cerebellar_Hemisphere
Brain_Caudate_basal_ganglia
Brain_Putamen_basal_ganglia
Brain_Amygdala
Brain_Hypothalamus
Brain_Substantia_nigra
Brain_Hippocampus

**T1D**

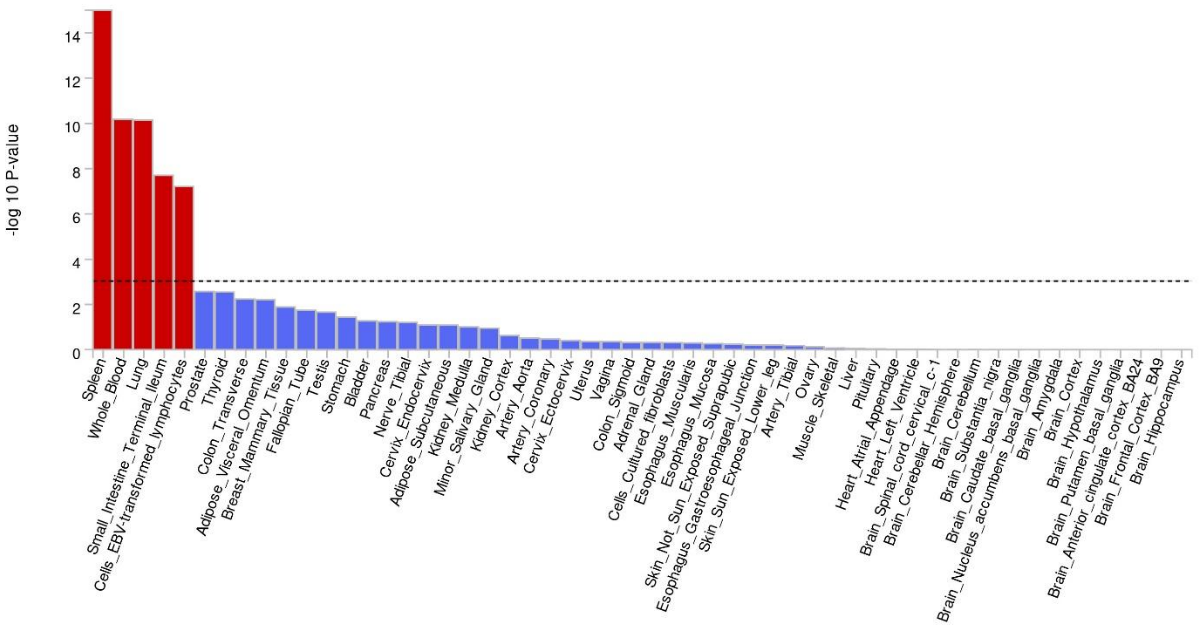

-log 10 P-value
Spleen
Whole_Blood
Lung
Small_Intestine_Terminal_Ileum
Cells_EBV-transformed_lymphocytes
Prostate
Thyroid
Colon_Transverse
Adipose_Visceral_Omentum
Breast_Mammary_Tissue
Fallopian_Tube
Testis
Stomach
Bladder
Pancreas
Nerve_Tibial
Cervix_Endocervix
Adipose_Subcutaneous
Kidney_Medulla
Minor_Salivary_Gland
Kidney_Cortex
Artery_Aorta
Artery_Coronary
Cervix_Ectocervix
Uterus
Vagina
Colon_Sigmoid
Adrenal_Gland
Cells_Cultured_fibroblasts
Esophagus_Muscularis
Esophagus_Mucosa
Skin_Not_Sun_Exposed_Suprapubic
Esophagus_Gastroesophageal_Junction
Skin_Sun_Exposed_Lower_leg
Artery_Tibial
Ovary
Muscle_Skeletal
Liver
Pituitary
Heart_Atrial_Appendage
Heart_Left_Ventricle
Brain_Spinal_cord_cervical_c-1
Brain_Cerebellar_Hemisphere
Brain_Cerebellum
Brain_Substantia_nigra
Brain_Caudate_basal_ganglia
Brain_Nucleus_accumbens_basal_ganglia
Brain_Amygdala
Brain_Cortex
Brain_Hypothalamus
Brain_Putamen_basal_ganglia
Brain_Anterior_cingulate_cortex_BA24
Brain_Frontal_Cortex_BA9
Brain_Hippocampus


**CD**

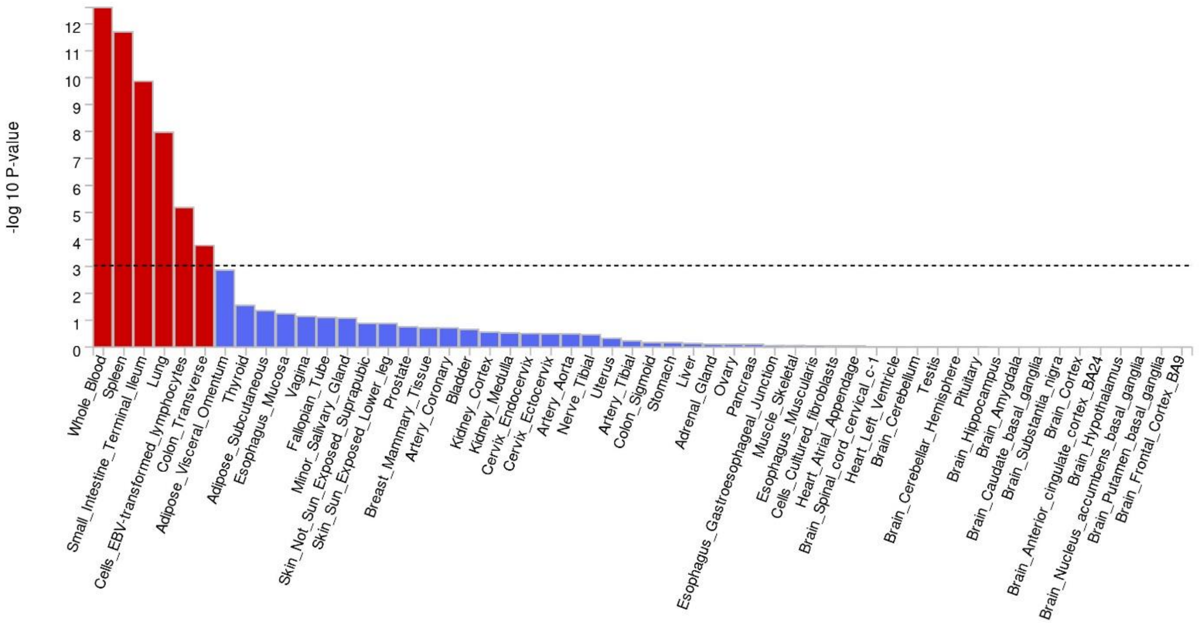

-log 10 P-value
Whole_Blood
Spleen
Small_Intestine_Terminal_Ileum
Lung
Cells_EBV-transformed_lymphocytes
Colon_Transverse
Adipose_Visceral_Omentum
Thyroid
Adipose_Subcutaneous
Esophagus_Mucosa
Vagina
Fallopian_Tube
Minor_Salivary_Gland
Skin_Not_Sun_Exposed_Suprapubic
Skin_Sun_Exposed_Lower_leg
Prostate
Breast_Mammary_Tissue
Artery_Coronary
Bladder
Kidney_Cortex
Kidney_Medulla
Cervix_Endocervix
Cervix_Ectocervix
Artery_Aorta
Nerve_Tibial
Uterus
Artery_Tibial
Colon_Sigmoid
Stomach
Liver
Adrenal_Gland
Ovary
Pancreas
Esophagus_Gastroesophageal_Junction
Muscle_Skeletal
Esophagus_Muscularis
Cells_Cultured_fibroblasts
Heart_Atrial_Appendage
Brain_Spinal_cord_cervical_c-1
Heart_Left_Ventricle
Brain_Cerebellum
Testis
Brain_Cerebellar_Hemisphere
Pituitary
Brain_Hippocampus
Brain_Amygdala
Brain_Caudate_basal_ganglia
Brain_Substantia_nigra
Brain_Cortex
Brain_Anterior_cingulate_cortex_BA24
Brain_Hypothalamus
Brain_Nucleus_accumbens_basal_ganglia
Brain_Putamen_basal_ganglia
Brain_Frontal_Cortex_BA9

**PSC**

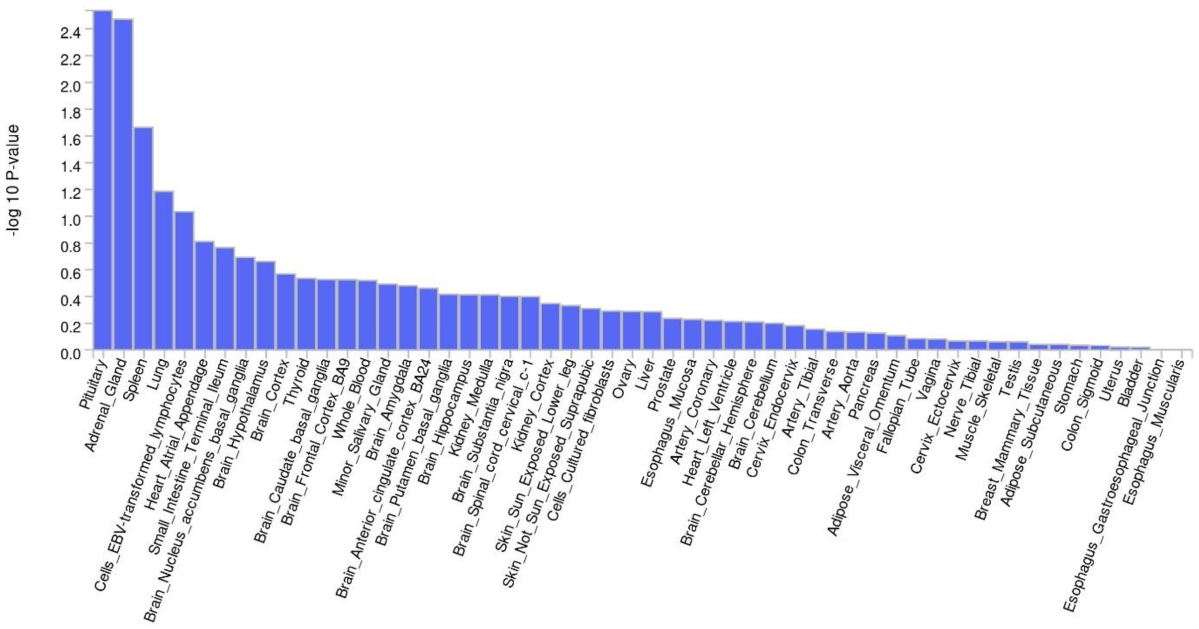
-log 10 P-value
0.0
0.2
0.4
0.6
0.8
1.0
1.2
1.4
1.6
1.8
2.0
2.2
2.4
Pituitary
Adrenal_Gland
Spleen
Lung
Cells_EBV-transformed_lymphocytes
Heart_Atrial_Appendage
Small_Intestine_Terminal_Ileum
Brain_Nucleus_accumbens_basal_ganglia
Brain_Hypothalamus
Brain_Cortex
Thyroid
Brain_Caudate_basal_ganglia
Brain_Frontal_Cortex_BA9
Whole_Blood
Minor_Salivary_Gland
Brain_Amygdala
Brain_Anterior_cingulate_cortex_BA24
Brain_Putamen_basal_ganglia
Brain_Hippocampus
Kidney_Medulla
Brain_Substantia_nigra
Brain_Spinal_cord_cervical_c-1
Kidney_Cortex
Skin_Sun_Exposed_Lower_leg
Skin_Not_Sun_Exposed_Suprapubic
Cells_Cultured_fibroblasts
Ovary
Liver
Prostate
Esophagus_Mucosa
Artery_Coronary
Heart_Left_Ventricle
Brain_Cerebellar_Hemisphere
Brain_Cerebellum
Cervix_Endocervix
Artery_Tibial
Colon_Transverse
Artery_Aorta
Pancreas
Adipose_Visceral_Omentum
Fallopian_Tube
Vagina
Cervix_Ectocervix
Nerve_Tibial
Muscle_Skeletal
Testis
Breast_Mammary_Tissue
Adipose_Subcutaneous
Stomach
Colon_Sigmoid
Uterus
Bladder
Esophagus_Gastroesophageal_Junction
Esophagus_Muscularis

**UC**

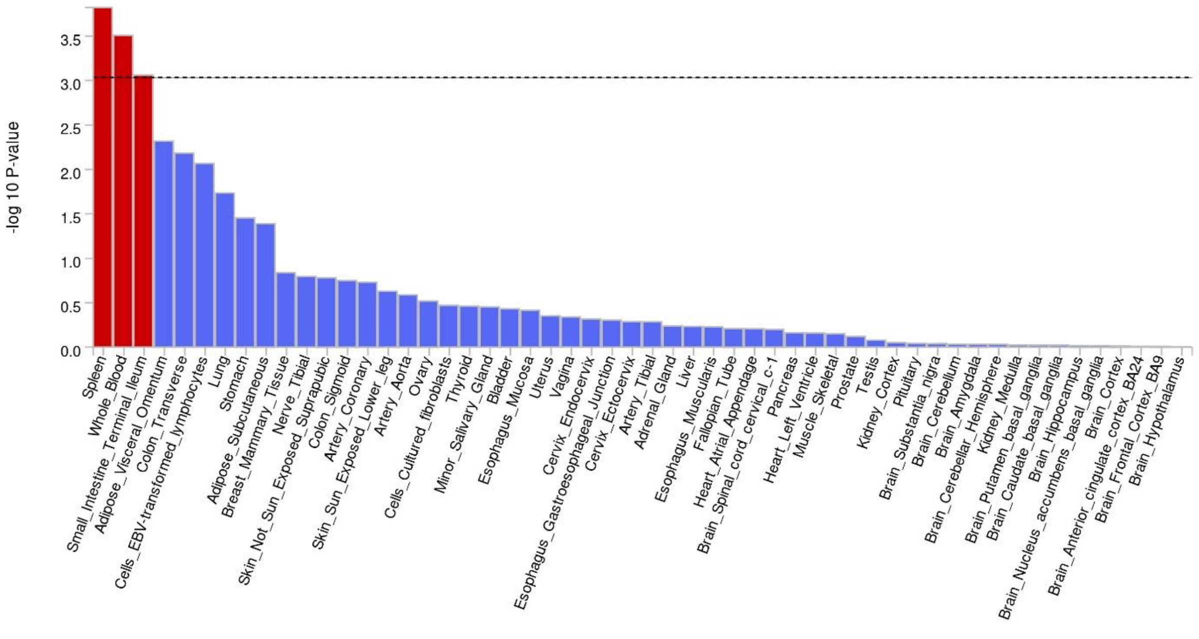
-log 10 P-value
0.0
0.5
1.0
1.5
2.0
2.5
3.0
3.5
Spleen
Whole_Blood
Small_Intestine_Terminal_Ileum
Adipose_Visceral_Omentum
Colon_Transverse
Cells_EBV-transformed_lymphocytes
Lung
Stomach
Adipose_Subcutaneous
Breast_Mammary_Tissue
Nerve_Tibial
Skin_Not_Sun_Exposed_Suprapubic
Colon_Sigmoid
Artery_Coronary
Skin_Sun_Exposed_Lower_leg
Artery_Aorta
Ovary
Cells_Cultured_fibroblasts
Thyroid
Minor_Salivary_Gland
Bladder
Esophagus_Mucosa
Uterus
Vagina
Cervix_Endocervix
Esophagus_Gastroesophageal_Junction
Cervix_Ectocervix
Artery_Tibial
Adrenal_Gland
Liver
Esophagus_Muscularis
Fallopian_Tube
Heart_Atrial_Appendage
Brain_Spinal_cord_cervical_c-1
Pancreas
Heart_Left_Ventricle
Muscle_Skeletal
Prostate
Testis
Kidney_Cortex
Pituitary
Brain_Substantia_nigra
Brain_Cerebellum
Brain_Amygdala
Brain_Cerebellar_Hemisphere
Kidney_Medulla
Brain_Putamen_basal_ganglia
Brain_Caudate_basal_ganglia
Brain_Hippocampus
Brain_Nucleus_accumbens_basal_ganglia
Brain_Cortex
Brain_Anterior_cingulate_cortex_BA24
Brain_Frontal_Cortex_BA9
Brain_Hypothalamus

**MS**

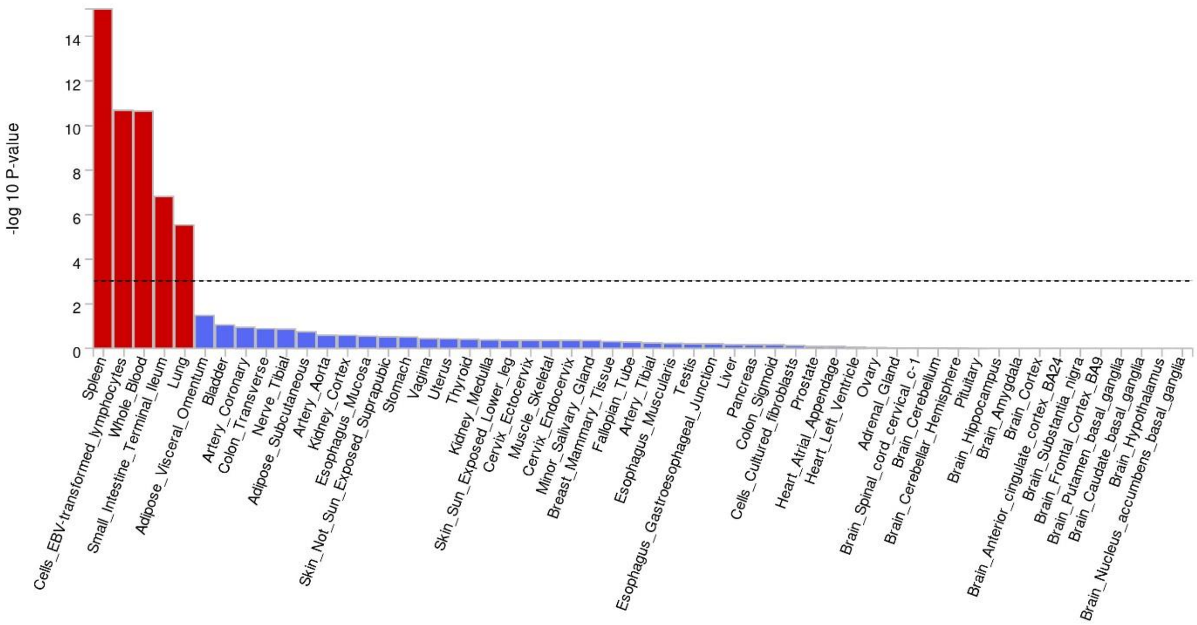

-log 10 P-value
0
2
4
6
8
10
12
14
Spleen
Cells_EBV-transformed_lymphocytes
Whole_Blood
Small_Intestine_Terminal_Ileum
Lung
Adipose_Visceral_Omentum
Bladder
Artery_Coronary
Colon_Transverse
Nerve_Tibial
Adipose_Subcutaneous
Artery_Aorta
Kidney_Cortex
Esophagus_Mucosa
Skin_Not_Sun_Exposed_Suprapubic
Stomach
Vagina
Uterus
Thyroid
Kidney_Medulla
Skin_Sun_Exposed_Lower_leg
Cervix_Ectocervix
Muscle_Skeletal
Cervix_Endocervix
Minor_Salivary_Gland
Breast_Mammary_Tissue
Fallopian_Tube
Artery_Tibial
Esophagus_Muscularis
Testis
Esophagus_Gastroesophageal_Junction
Liver
Pancreas
Colon_Sigmoid
Cells_Cultured_fibroblasts
Prostate
Heart_Atrial_Appendage
Heart_Left_Ventricle
Ovary
Adrenal_Gland
Brain_Spinal_cord_cervical_c-1
Brain_Cerebellum
Brain_Cerebellar_Hemisphere
Pituitary
Brain_Hippocampus
Brain_Amygdala
Brain_Cortex
Brain_Anterior_cingulate_cortex_BA24
Brain_Substantia_nigra
Brain_Frontal_Cortex_BA9
Brain_Putamen_basal_ganglia
Brain_Caudate_basal_ganglia
Brain_Hypothalamus
Brain_Nucleus_accumbens_basal_ganglia

**Supplementary 2., Fig. 6. MAGMA-identified genes uniquely associated with diseases involved in factor 4 and linked to monogenic immune disorders and blood syndrome**. Janus kinase 2 is involved in different blood disorders, *KLHDC8B* is involved in Hodgkin lymphoma, *RTEL1* in inborn error failure. *IRF1* and *CARD9* engage in different types of immunodeficiencies [Rosain et al., 2023, Wang et al., 2019, Salipante et al., 2009]. The *NOD2* gene plays a role in Blau syndrome, a rare autosomal dominant autoinflammatory syndrome classified as an autoinflammatory phenotype according to Tangye et al., 2022. Also, *STAT5B* gene is the monogenic cause of autosomal dominant growth hormone insensitivity syndrome with immune dysregulation-2.

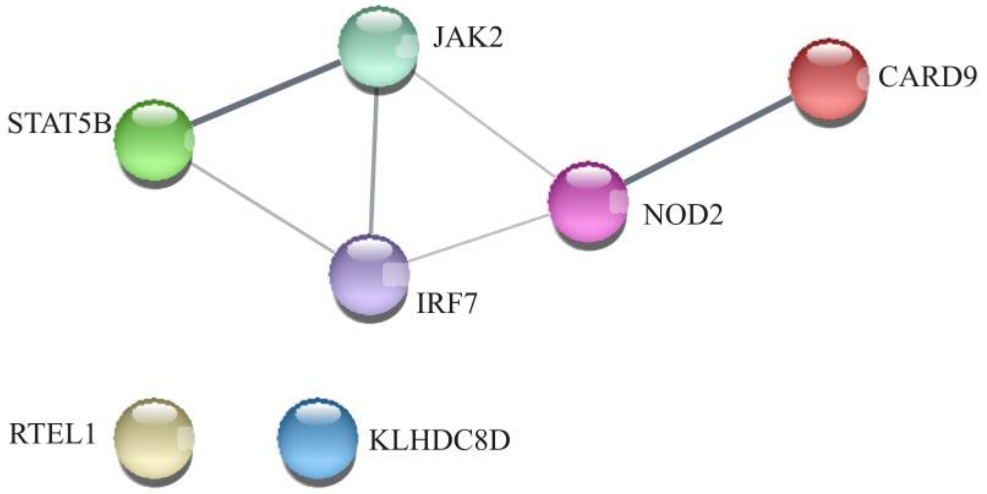
